# Analysis of electronic health records from three distinct and large populations reveals high prevalence and biases in the co-administration of drugs known to interact

**DOI:** 10.1101/2023.02.06.23285566

**Authors:** Jon Sánchez-Valle, Rion Brattig Correia, Marta Camacho-Artacho, Rosalba Lepore, Mauro M. Mattos, Luis M. Rocha, Alfonso Valencia

## Abstract

The co-administration of drugs known to interact has a high impact on morbidity, mortality, and health economics. We study the drug-drug interaction (DDI) phenomenon by analyzing drug administrations from population-wide Electronic Health Records (EHR) in Blumenau (Brazil), Catalonia (Spain), and Indianapolis (USA). Despite very different health care systems and drug availability, we find a common large risk of DDI administration that affected 13 to 20% of patients in these populations. In addition, the increasing risk of DDI as patients age is very similar across all three populations but is not explained solely by higher co-administration rates in the elderly. We also find that women are at higher risk of DDI overall— except for men over 50 years old in Indianapolis. Finally, we show that PPI alternatives to Omeprazole can reduce the number of patients affected by known DDIs by up to 21% in both Blumenau and Catalonia, and 2% in Indianapolis, exemplifying how analysis of EHR data can lead to a significant reduction of DDI and its associated human and economic costs. Although the risk of DDIs increases with age, administration patterns point to a complex phenomenon that cannot be solely explained by polypharmacy and multimorbidity. The lack of safer drug alternatives, particularly for chronic conditions, further overburdens health systems, thus highlighting the need for disruptive drug research.

## 1. Introduction

Adverse drug reactions (ADR) are noxious or unintended effects related to drug administration. ADR are a major public health problem due to their impact on morbidity, mortality, and health economics [1, 2]. The co-administration of drugs may cause ADR from a drug-drug interaction (DDI), defined as the effect one drug has on another either at the pharmacokinetic or pharmacodynamic level. ADR have been associated with 4.2% to 8.4% of all hospital admissions [2, 3], and of these, about 51% are related to DDIs [2]. These numbers increase with polypharmacy. The risk of ADR-related hospital admission goes up from five-fold for patients treated with more than three drugs, to nine-fold for those treated with more than 10 drugs [2]. Because instances of polypharmacy increase due to higher prevalence of multimorbidity (the cooccurrence of two or more diseases in the same patient) with aging, so does the risk of DDIs [4, 5]. For instance, a study analyzing elderly outpatients in six European countries finds that 46% of outpatients had at least one potentially significant DDI, and 10% had severe interactions [6].

Factors in addition to age, such as patient sex [5, 7], errors and lack of information in ambulatory care [8, 9], and number of physicians prescribing drugs [10], are known to also increase the risk of DDIs. Often, physicians are unaware of the complete list of the drugs their patients are taking [9]. To counter this, computerized health information systems (HIS) such as Electronic Health Records (EHR), drug interaction software, and decision support systems have been developed to proactively screen for DDIs and alert physicians and pharmacists [11]. Even though reports of preventable ADR-related hospital admissions vary widely, from 24-52% [12, 13] to 77-92% of all ADR-related hospital admissions [2, 14], HIS attempt to lower these rates. However, HIS alone are not enough to prevent prescription errors, as physicians may simply dismiss alerts [15]. Together, these distinct factors paint a picture of a complex DDI phenomenon with worrying direct consequences for both patients and health systems. For instance, our own analysis revealed that DDIs likely account for a significant financial burden to public health, reaching 2 dollars per capita in a city in Brazil during an 18-month period—extrapolated to an expenditure of $565M for the country in the same period [5].

To better untangle the factors involved in the global DDI phenomenon, we analyze administration patterns retrieved from EHR from three large populations with distinct public and private healthcare systems: Blumenau (Brazil; pop. 338,876), Catalonia (Spain; pop. 7.6 million), and Indianapolis (USA; pop. 876,682). We study demographic variables, such as age and sex, as well as drugs involved in DDIs in all three populations in detail. In addition, we evaluate the role of polypharmacy and co-administration by building a statistical null-model that shuffles drug labels while accounting for cohort-specific drug availability. Finally, we demonstrate the population-level impact of individual DDIs by simulating administration of drug alternatives to Omeprazole, a commonly prescribed proton pump inhibitor with several known and avoidable interactions.

## 2. Results

### 2.1 Population comparison

In order to best compare the three populations, we first analyze the initial 18 months (the smallest temporal window available, for Blumenau) of administrations in each population. This is necessary as longer study periods increase the chances of observing co-administrations and DDIs and could bias our conclusions. We find that 140, 814, and 1,228 unique drugs were dispensed respectively in Blumenau, Catalonia, and Indianapolis, with 106 drugs common to all three populations (see fig. S4A). The three populations present a very similar risk of co-administration (*RC*) with the largest for Blumenau (76.99%), followed by Catalonia (75.78%), and Indianapolis (74.16%). This risk increases to 89.83% for Catalonia and 75.53% for Indianapolis when we analyze all available data (11 and 2 years, respectively; see table S2).

Given the common set of 106 drugs, we observe 149 known DDI pairs co-administered in all three populations (fig. S4B). The three populations also observe a similar risk of drug interaction (*RI*), with the largest again for Blumenau (12.51%), but closely followed by Indianapolis (12.12%), and then Catalonia (10.06%). This risk increases to 20.36% for Catalonia and 13.04% for Indianapolis when we analyze all available data (11 and 2 years, respectively; see table S2). Further leveraging all available data we show that the DDI phenomenon is more similar between Catalonia and Blumenau (0.52, Spearman correlation, see fig. S4C), in comparison to Indianapolis and either Catalonia (0.3) or Blumenau (0.27).

### 2.2 Sex risk comparison

We observe only a slightly higher but significant relative risk of co-administration for women in the three populations (*eq. 8*): Blumenau (*RRC*^*W*^ = 1.07), Indianapolis (*RRC*^*W*^ = 1.06), Catalonia (*RRC*^*W*^ = 1.05) (see table 1). This risk increases substantially when focused on interacting drugs, especially in Blumenau (*RRI*^*W*^ = 1.54), but is also high in Catalonia (*RRI*^*W*^ = 1.25), and present in Indianapolis (*RRI*^*W*^ = 1.12). Drug combinations that cause moderate interactions, which should be used only under special circumstances because of their clinically significant outcomes, are the most co-administered in all three populations, and drive the differences between sexes (see Table 1).

**Table 1.** Relative risk for women (RRW) of drug co-administration (RRCW) and interactions (RRIW); the latter is also computed for types of interactions as per drugs.com (minor, moderate, and major). The percentage of patients of each sex (M = man; W = woman) for each case is also shown. Values shown for all three populations during the first 18 months of the study. Asterisks denote statistically significant relative risks based on Fisher’s exact test results.

|  | RRW | Blumenau<br>% W - M | RRW | Catalonia<br>% W- M | RRW | Indianapolis<br>% W - M |
| --- | --- | --- | --- | --- | --- | --- |
| Co-administration ($RRC^W$) | 1.07* | (79.08% - 74.06%) | 1.05* | (77.39% - 73.88%) | 1.06* | (75.88% - 71.82%) |
| Interaction ($RRI^W$) | 1.54* | (14.64% - 9.49%) | 1.25* | (11.08% - 8.84%) | 1.12* | (12.69% - 11.36%) |
| Minor Interaction | 0.81* | (0.36% - 0.44%) | 1.27* | (0.26% - 0.2%) | 1.02 | (1.13% - 1.1%) |
| Moderate interaction | 1.59* | (11.3% - 7.1%) | 1.4* | (8.31% - 5.93%) | 1.12* | (10.67% - 9.49%) |
| Major interaction | 1.53* | (6.25% - 4.07%) | 1.08* | (3.17% - 2.95%) | 1.02 | (4.87% - 4.76%) |

### 2.3 Age risk comparison

To analyze the effect of patient ageing on the risk of drug co-administration and DDIs, we divide patients into age intervals of 5 years, based on their age at the time of administration (see section 4.4). As a well-known polypharmacy phenomenon, the risk of co-administration (*RC*^[*y*1,*y*2]^, *eq. 10*) increases with age in all three populations as depicted in fig. 1a; fig. S5 depicts the proportions of patients per number of drugs simultaneously co-administered. It is noteworthy that there is a drop in *RC* in the 10-14 age range for all three populations. Patients in the 15-59 year-old range in Catalonia have the lowest *RC*, although the largest *RC* is also observed in Catalonia for patients older than 59. Conversely, it is in Indianapolis that the largest *RC* is observed for 20-59 year-old patients.

**Figure 1.**
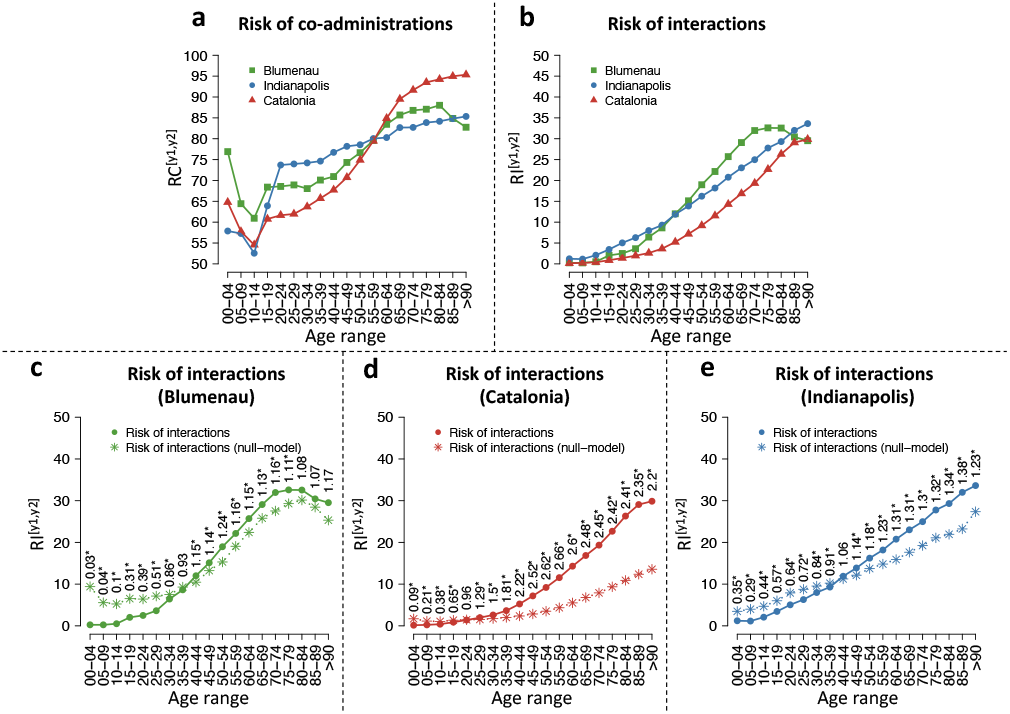
Risk of co-administration and interaction by age during the first 18 months of the studies. Green, red, and blue lines denote measurements for Blumenau, Catalonia, and Indianapolis, respectively. (a) Risk of co-administration of drugs, *RC*^[*y*1,*y*2]^ (*eq. 10*). (b) Risk of co-administration of drugs known to interact, *RI*^[*y*1,*y*2]^ (*eq. 11*). (c-e) *RI*^[*y*1,*y*2]^ against respective null model 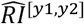 in (c) Blumenau, (d) Catalonia, and (e) Indianapolis. Circles denote the values obtained with the real data, while the asterisks denote the values obtained using the null model. The associated relative risk (*eq. 13*) is shown above the points. Asterisks denote significant differences (Fisher’s exact test).

The risk of a DDI (*RI*^[*y*1,*y*2]^, *eq. 11*) increases with age from less than 0.2% of patients in the 0-4 year range, to up to 33.6% of patients over 90 years-old (see fig. 1b). After the age of 75, *RI* is at least 20% for all three populations, and over 32% for Blumenau. Interestingly, all three populations display monotonically increasing *RI* with age (except for the oldest two age groups in Blumenau), despite their widely different cultures, available medications, and health care systems. Despite this, there are some noteworthy differences among the three populations as well. For instance, Indianapolis has the highest *RI* in patients age 0-39 as well as those older than 85. Blumenau, on the other hand, has the highest *RI* for patients age 40-84, being Catalonia the one with the lowest *RI* across all age groups, even though its patients age 60-90 have the highest *RC* (compare fig. 1a&b).

To evaluate whether the observed increasing *RI* with age in all three populations (fig. 1b) is explained by the also increasing *RC*, we build a statistical null model, marked in fig. 1c-e with asterisks, which yields the expected 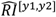 for each age range [*y*1,*y*2] if patients were prescribed (age-specific) drugs at random. Random prescription of drugs is of course oblivious to know DDI information, so one would expect actual prescription—given available information about DDI—to result in lower risk than the null model. Indeed this is observed for younger age groups, as the actual *RI* is lower than that of the null model with random drug administration. Thus, younger patients are at a lower-than-random risk of DDIs for their rate of drug co-administration. However, and much to our surprise, for patients over 20 years of age in Catalonia and over 40 years of age in Blumenau and Indianapolis, the actual *RI* significantly surpasses what would be expected by chance: a worse-thanrandom chance of administering DDIs. This means that the higher risk of drug interactions faced by older age groups cannot be explained solely by increasing polypharmacy.

### 2.4 Sex risk by age comparison

To study the role of sex in the observed age-associated risk of co-administration and DDI during the first 18 months of data in all three populations, we also analyze men and women separately. Figure 2a-c shows that women consistently have a higher risk of drug co-administration (*RRC*^*W*^, *eq. 8*) throughout their lifetime in all three populations, when compared to men. Nonetheless, this relative risk is typically small, being significant in almost all age ranges in Catalonia and only in specific age ranges in the cases of Indianapolis and Blumenau (15-29 years old). Overall, in Catalonia, we observe the smallest *RRC*^*W*^ across all ages, with greater sex imbalance in co-administration observed in Blumenau and Indianapolis showing; across most age groups in the former, and greater imbalance for women only in age group 15-44 in the latter.

**Figure 2.**
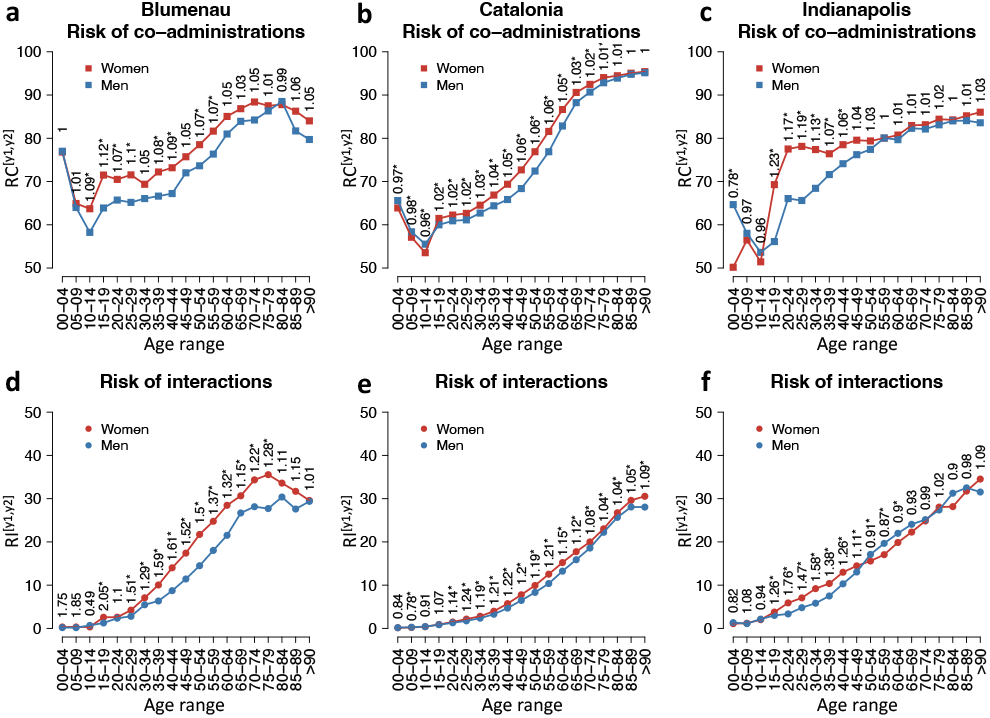
(a-c) Risk of drug co-administration, *RI*^[*y*1,*y*2],*g*^, and (d-f) interaction, *RI*^[*y*1,*y*2],*g*^, by age and sex (as defined in section 4.6) for Blumenau, Catalonia, and Indianapolis in the first 18 months of administration. Red and blue colors denote the risks in women (*g* = *W*) and men (*g* = *M*), respectively. Relative risks of co-administration (*RRI*^[*y*1,*y*2],*W*^) and interaction (*RRI*^[*y*1,*y*2],*W*^) for women per age group displayed above the points (as defined in section 4.5 and section 4.6). Asterisks denote significant differences (Fisher’s exact test).

The cross population comparison of the risk of sex-related drug interaction (*RRI*^*W*^, *eq. 9*) across age groups, reveals some similarities as well as more nuanced differences. *RRI*^*W*^ is higher for women in Blumenau and Catalonia in almost all age ranges, with the exception of younger age groups (10-14 in Blumenau and 0-14 in Catalonia as shown in fig. 2d-e). In contrast, in Indianapolis, men are at a higher risk of DDI in the 50-89 age range, significantly so for patients aged 50-64, as seen in fig. 2f. Nonetheless, the relative risk of interaction reaches higher values for women than for men in all three populations. In Catalonia, which presents the most sex-balanced scenario across age groups, women aged 25-59 face a significant risk of interaction in comparison to men near or above 20% (*RRI*^[25,59],*W*^ ≥ 1.19). Interestingly, when we analyze all 11 years of data for Catalonia, the risk for younger women is also above 20% with *RRI*^[15,59],*W*^ ≥ 1.2 (see fig. S6e). In fact, when analyzing all 11 years worth of data, the largest relative risk of DDI for women is observed in the 15-29 age range, which correlates with higher Ethinylestradiol administrations in the year 2012 (fig. S7a and fig. S8d-e).

In Indianapolis, women aged 15-44 face a risk of interaction at least 26% higher than men (*RRI*^[15,44],*W*^ ≥ 1.26), peaking at *RRI*^[20,24],*W*^ = 1.76. In Blumenau, women aged 25-64 face a risk of interaction in comparison to men near or above 30% (*RRI*^[25,64],*W*^ ≥ 1.29), reaching a peak at *RRI*^[40,44],*W*^ = 1.61. In summary, across the three populations, women between 15 and 49 face a substantially higher risk than men of administering a known DDI—the largest risk is observed in Blumenau for women aged 15-19 (*RRI*^[15,19],*W*^ = 2.05). When compared to the null model 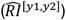, we note that the worst-than-random risk of interaction happens earlier for Catalan women (15-19 age range) than for men (20-24) (fig. S9). For Blumenau and Indianapolis there is no sex difference when comparing to the null model.

Naturally, DDIs can cause different levels of adverse events, from mild headaches to patient hospitalization due to liver damage complications. Thus, we study the sex-associated differences based on the severity of the DDI, by tallying the number of women and men in each age range while accounting for minor, moderate, and major DDIs. DDI severity is extracted from drugs.com [16] (see section 4.4). Results are shown in figs. S10 and S11 and indicate that moderate DDIs are the most common with increasing patient age. In addition, in Indianapolis, the shift in gender-associated risk is largely explained by moderate DDIs, more common in women 15-49 years old and in men over 50 (see fig. S10j). An interesting pattern of elevated risk for major DDIs in older men is also present in both Catalonia and Indianapolis, but not Blumenau. In Catalonia, men have a higher risk of major DDIs in the ages 50-84 (see fig. S10g, while in Indianapolis men have a higher risk of major DDIs in ages 45-84 (see fig. S10k). Since drugs.com is tailored to an U.S. audience, drugs administered in other countries and their associated interactions may not be included in the site. The differences in the risk of administering these DDI is very similar in the three populations, being higher for women in Blumenau, and for men in Catalonia and Indianapolis.

### 2.5 Drug interaction networks

To better characterize the DDI phenomenon in each of the three populations, as described in section 4.7, we build drug-drug interaction networks shown in fig. 3 and figs. S1 to S3.

**Figure 3.**
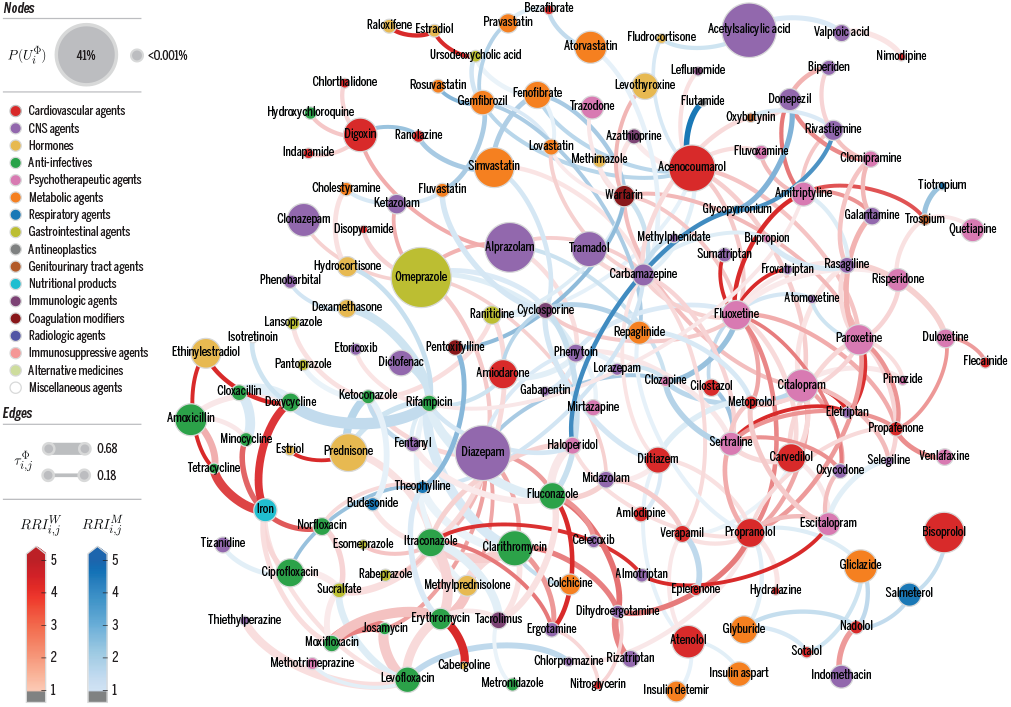
Catalonia DDI Network. Nodes denote drugs *i* involved in at least one co-administration known to be a DDI. Only nodes connected via edges with 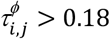 are shown for clarity. Node color represents the highest level of primary action class, as retrieved from drugs.com. Node size proportional to 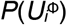 per *eq. 12*, the probability of patients being affected by a DDI involving drug *i*. Edge weights denote strength of interaction, 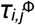 per *eq. 5*. Edge colors denote 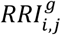, where *g* ∈ *M, W*, to identify DDI edges that are higher risk for women (red) or men (blue). Color intensity for 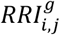 varies in [1,5]; that is, values are clipped at 5 for clarity.

Nodes are colored based on their drugs.com category and sized based on the probability that patients prescribed the drug will experience a DDI (*eq. 12*). Edge width represents the strength of drug interaction (*eq. 6*) and edge color denotes the sex-associated relative risk of a DDI (*eq. 9*), with red (blue) denoting higher risk for women (men). An interactive version of these networks can be explored at http://disease-perception.bsc.es/ddinteract/.

These networks help us visualize not only which drugs are most involved in interactions but also to identify pairs with the same sex-associated bias (edge color) in all populations. For instance, considering the 149 DDIs common to all three populations, 56% are associated with increased risk for the same sex (56 DDIs for women, 27 for men). In addition, the network representation facilitates inferences for specific drugs or categories. For instance, drug pairs associated with Fluconazole, contraceptives, or benzodiazepines tend to be associated with higher risk for women, while most of the interactions associated with anticoagulants (Warfarin with Phenytoin, Prednisone, Amiodarone, etc) represent higher risk for men.

Conversely, there are drug pairs where the sex-associated bias is reversed in at least one population, with Blumenau presenting the highest discordance: 27 pairs. Interestingly, 11 of these 27 discordant interactions are major DDIs, including the concomitant use of ASA (anticoagulant) and Ibuprofen (anti-inflammatory), a combination that reduces the effectiveness of aspirin in preventing stroke and increases the risk of developing gastrointestinal ulcers (see table S5).

### 2.6 Drug interactions driving sex-associated biases

Among the shared drug interactions in all three populations (149), we observe a strong association between Omeprazole and both Clonazepam and Diazepam for women in Blumenau and Catalonia (see red cells in fig. 4ab), but not in Indianapolis (see fig. S12). This is particularly supported by the over administration of Omeprazole in the two populations (see table S9). Similarly, the risk of co-administering Alendronic acid—used to treat osteoporosis—and nonsteroidal anti-inflammatories is higher for women, paired with Diclofenac in Catalonia and Ibuprofen in both Blumenau and Catalonia. This DDI may result in increased risk for stomach and intestine irritation. The co-administration of Ethinylestradiol (contraceptive) and Amoxicillin (antibiotic) is significantly high in all three populations. This DDI may result in reduced contraceptive effectiveness, thus increasing the risk of unwanted pregnancy. Interestingly, the major interaction between ASA and Ibuprofen previously observed to be associated with an higher risk for women in Blumenau [5], is conversely associated with a lower risk for women in the other two populations (see figs. 4 and S12), suggesting a particularity of the Blumenau health care system. This result points to the existence of cultural or social factors that play a role in this sex-associated bias. Another interesting DDI case that may point to social or cultural factors is the drug pair Lidocaine-Carvedilol, that only presents a higher risk for men in Indianapolis (see fig. S12).

**Figure 4.**
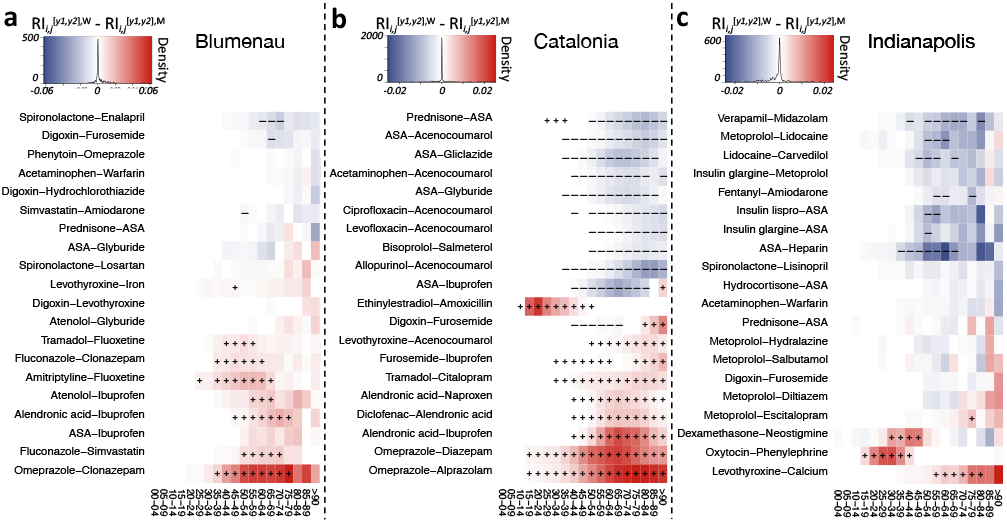
Top 20 drug interactions with the highest difference between *RI*^[*y*1,*y*2],*W*^ and *RI*^[*y*1,*y*2],*M*^ (see *eq. 11*). Colors denote a higher risk of interaction for women (red) and men (blue). Markers (+ and −) denote significantly higher risk of DDI administration for the respective sex after correcting for multiple testing (FDR ≤ 0.05). Note color scale is different across populations, as the maximum and minimum differences in *RI*^[*y*1,*y*2]^ are different between populations.

Looking further at the DDIs with high sex-and age-associated risks in each population (Figure 4), we notice in Blumenau a significantly higher risk for women of co-administering Fluoxetine (major depression treatment) with Tramadol (opioid analgesic) or Amitriptyline (tricyclic antidepressant). In Catalonia, for men over 40 years old, the risk of co-administering anticoagulants such as ASA and Acenocoumarol either with each other or with anti-diabetic drugs (Gliclazide and Glyburide), Allopurinol (gout treatment), Prednisone (glucocorticoid anti-inflammatory), or antibiotics (Ciprofloxacin and Levofloxacin) is significantly higher (see fig. 4b). Lastly, in Indianapolis young women are at a significantly higher risk of co-administering Oxytocin, used to induce labor, and Phenylephrine, used to increase blood pressure (see fig. 4c). For women older than 55 in Indianapolis, there is also a significantly higher risk of co-administering Levothyroxine, used to treat hypothyroidism, with Calcium, which can change the absorption levels of Levothyroxine. Conversely, we also found drug pairs with an increased risk for men. For instance, the combination of two anticoagulants, ASA and Heparin; Verapamil (a calcium channel blocker) and Midazolam (benzodiazepine); Lidocaine (a local anesthetic) with Metoprolol and Carvedilol (a beta blocking agent); and anti-diabetic drugs, such as Insulin lispro and Insulin glargine, with ASA and Metoprolol, a beta1 receptor blocker used to treat high blood pressure that can increase the risk of hypoglycemia. Specific interacting pairs can be visualized at http://disease-perception.bsc.es/ddinteract/

### 2.7 Alternative drug treatments to avoid DDIs

While the observed DDIs involving Omeprazole and either Clonazepam or Diazepam are mostly irrelevant in Indianapolis (administered by 256 and 135 patients, respectively), they are the most co-administered drug pairs in Blumenau (5,076, 998) and Catalonia (47,811, 253,473). Here, we analyze the preferential co-administration of Omeprazole over alternative proton-pump inhibitors (PPI) that have no known drug-interaction with benzodiazepines in Catalonia (see section 4.9). Catalonia presents a significant preferential co-administration of Omeprazole with Diazepam or Clonazepam, as compared to other PPI as a group (i.e., Esomeprazole, Pantoprazole, Rabeprazole and Lansoprazole) (OR = 17.6 and 12.2, respectively) or individually (see table S6). Conversely, in Indianapolis, there is a significant preferential administration of alternative PPI in combination with Diazepam or Clonazepam (OR=38.3 and 13.5). Importantly, alternative PPI are available for administration in Catalonia, which is not the case for the public health care system of Blumenau where they can only be purchased from private pharmacies. Indeed, 12 of the 16 (75%) drugs associated with Omeprazole interactions can be avoided using an alternative PPI.

Based in this observation, we first simulate for Catalonia the population-level effect of removing the Omeprazole-associated interactions from the overall DDI risk. In this simulation, we replace Omeprazole with currently available alternative PPI and recalculate the DDI risk. We find that administering alternative PPI reduces the overall levels of DDI in Catalonia by 23.28% in women and 20.09% in men (see fig. S16b). The majority of these avoidable Omeprazole interactions are generating moderate adverse effects (see fig. S13b), which affect 18.85% (12.31%) of men (women) and can be avoided in 34.82% and 32.9% of the patients. For Indianapolis, the same simulation only reduces overall DDI levels by 2.55% in men and 2.56% in women (see fig. S16g-h). Though no Omeprazole substitutes are available free of charge in Blumenau, we followed the same simulation procedure using the alternatives available in Catalonia. Interestingly, the percentages of preventable interactions are almost identical to those in Catalonia, 23.19% for women and 19.51% for men (see fig. S16a-c).

## 3. Discussion

This is the first study to analyze DDI administration patterns in three large populations with distinct health care systems. In total, we analyze drug administration records from almost six million patients from up to 11 years worth of data. Despite different study periods and data resolutions for each population, similar patterns were revealed. The risk of drug co-administrations and interactions by age are both similar for the three populations (fig. 1ab). This shows that the DDI phenomenon is a public health burden in both developed and developing nations regardless of access to medication or type of health care system. Furthermore, our statistical null model, designed to account for polypharmacy while preserving the same number of prescribed drugs and co-administrations per age, shows that the much higher risk of DDI in older age (in all populations) is not solely explained by higher risk of co-administration in those age groups. Indeed, this worrisome result previously observed in Blumenau [5], is here shown to be even worse in Catalonia, where patients have a worse-than-random risk of DDI starting early in their twenties—reaching 2.7 fold higher-than-random risk for 55 to 59 year-olds (see fig. 1d). This worse-than-random risk of DDI remains even when separating men and women populations (see fig. S9), questioning multimorbidity treatments and its current focus on geriatric patients.

Also similarly observed in all populations is a higher risk for women of both drug co-administration and interactions in comparison to men. The general risk of co-administration for women increases as they age, although the largest difference from men occurs during peak reproductive age (age ranges 15-29, see fig. 2 and fig. S6) which may be explained by the greater use of the healthcare systems by women during these years [17]. On the other hand, the sex imbalance risk is generally much higher for interactions than for co-administration (see fig. S17). There are possible explanations as for why women have a generally higher risk of DDI. For instance, some drugs are simply women-specific, such as hormones and contraceptive drugs. Thus, women-specific drugs may partially explain the higher risk of DDI observed, particularly for younger women. The DDI pair Ethinylestradiol and Amoxicillin was jointly given to 0.98% of Catalan women, but only to 0.0008% of men. In Blumenau, this same drug pair was given to 0.6% of women, and to no men. Unfortunately, we cannot infer from our data whether prescribers informed the patients of this DDI and potential need for additional contraceptive methods during co-administration.

Additional reasons for the generally observed higher risk for women come from the fact that some diseases are more likely to affect women. For instance, osteoporosis is a skeletal disorder characterized by compromised bone strength [18] and known to be diagnosed more frequently in women [19]. This sex-associated prevalence is clearly observed in our data for the populations with disease codes (Catalonia and Indianapolis, see fig. S15). Bisphosphonates, such as Alendronic acid, are used to treat osteoporosis and, as a consequence, the risk of DDI related to Alendronic acid is higher for women, especially those over 50. For instance, the risk of interaction for women aged 60-64 between this drug and Ibuprofen is 1.8 and 1.34 for Catalonia and Blumenau, respectively. For men at the same age this risk is only 0.1 and 0.22 in both populations (see table S7). The same can be seen in Indianapolis albeit at a smaller scale. The risk of interaction for Alendronic acid and Ibuprofen is only 0.04 for women in Indianapolis at the same 60-64 age range. And virtually no men administered this DDI in Indianapolis (see table S7). This smaller risk for Indianapolis is further supported by the comparatively small administration of *Alendronic acid* (0.5% compared to 3.5% and 1.7%, see table S8) which likely stems from the decreased use of bisphosphonates in the US after the 2010 FDA bisphosphonate drug safety communication [20].

A deviation from the general trend of increased DDI risk for women is particularly noteworthy. In Indianapolis, men over 50 years of age do have a higher risk of DDI than women. This difference is driven by two factors. First, less frequent use of Omeprazole in combination with benzodiazepines, widely used for women in the other two populations and correlated with significantly higher risk there (see table S3). Indeed, when we remove Omeprazole administration in Catalonia from our analysis (see section 2.7), men over 60 also show higher risk of DDI than women (fig. S14). Second, the administration of some particular DDI that are given significantly more to men in Indianapolis, such as Verapamil-Midazolam, Metoprolol-Lidocaine, and Lidocaine-Carvedilol (see fig. 4c). These observations highlights how our study also reveals specifics of sex-related bias in the DDI phenomenon for each population. Given the tools we provide for further analysis, other researchers interested in this problem can further study and characterize specific DDIs of interest.

Another facet of the complex DDI phenomenon is patient multimorbidity. The proportion of patients with multimorbidities increases substantially with age, with almost 80% of the people suffering from at least two morbidities at the age of 65 [21]. As classical treatments are disease independent, patients with multimorbidities are particularly at increased risk for DDI. For instance, patients with type 2 diabetes are known to be at higher risk for cardiovascular diseases and thrombotic complications [22]. To treat both conditions, antidiabetic drugs such as Glyburide, Gliclazide, Insulin lispro, and Insulin glargine are often combined with anticoagulants such as ASA and Acenocoumarol (the last being dispensed only in our Catalonia data), which increases the risk of hypoglycemia. Our work highlights these are among the top 10 DDIs ranked by the number of patients they affect in all three populations. In addition, several of these drugs are usually co-administered for long periods of time, as characterized by our strength of interaction measure (see table S3). Also related to anticoagulants, gout, an inflammatory disease characterized by elevated levels of uric acid, is known to increase the risk of thrombosis [23]. As a potential consequence, we find a higher than expected chance of concomitantly prescribing Allopurinol with Warfarin (see table S1), a DDI that increases the risk of bleeding due to the potentiation of the anticoagulant effect [24]. Interestingly, the incidence of type 2 diabetes, and gout are higher for men over 50 in Catalonia (fig. S15) and can potentially explain the higher administration of the above mentioned DDIs.

An important aspect of our study is to exemplify how our large-scale study of the DDI phenomenon can lead to actionable interventions for public health benefit. For that purpose, we studied the role of the proton pump inhibitor Omeprazole on the observed DDIs in the three populations. PPI are the leading therapy for upper gastrointestinal disorders and prevention of gastric ulcers associated with the use of non-steroidal anti-inflammatories [25]. However, there is substantial evidence for inappropriate over-prescription of PPI, particularly of Omeprazole [26–29]. For instance, in 2008 it was estimated that 100 million pounds from the National Health Service budget, and almost 2 billion pounds worldwide, were being spent unnecessarily on PPI [28]. And four fifths of all PPI administrations in the UK were associated with Omeprazole.

The lack of awareness, overuse, and misuse of PPI, together with the elevated number of drug interactions associated with Omeprazole (Phenytoin, Methotrexate, and several benzodiazepine derivatives, among others), makes Omeprazole one of the most important culprits of DDIs. Indeed, in our study, Omeprazole is the 3^rd^ and 4^th^ most dispensed drug in Blumenau and Catalonia, respectively. Conversely, in Indianapolis it is the 44^th^. Therefore, we simulated the substitution of Omeprazole with alternative PPI—such as Pantoprazole and Lansoprazole––as a possible, but actionable, public health intervention. Such an intervention would result in a reduction of 20% of all men and 23% of all women currently administering a DDI in Catalonia (fig. S16b). This means 156,210 women and 92,533 men would be DDI-free in Catalonia if their Omeprazole prescription was substituted by another PPI. To put this in perspective, assuming 10% [6] of these patients had to seek hospital care due to this DDI, this intervention would amount to a total savings of 42 million euros for the Catalan health care system (calculated based on the average hospitalization cost of 1,709.85 euros for Catalonia in 2020 [30]). In contrast, extending the simulation to Indianapolis, results in a much smaller reduction of DDI risk (only 2.5% fewer patients would have not been administered a DDI, see fig. S16c). This shows that in Indianapolis the availability of PPI alternatives is being utilized to avoid known DDIs or ADR involving this drug. Thus, as actionable interventions, our study suggests that Catalonia should encourage prescription of available PPI alternatives.

Some limitations of our study are warranted. First, we assume that drugs dispensed were administered for their complete treatment length. In reality, patients may stop administration mid treatment, and prescribers may substitute drugs for patients with complaints of adverse effects. Also, adverse drug reactions may in some cases be avoided by separating drug intake during the day or adjusting dosage. Thus, our results should be seen as a worst-case scenario for the administration of known DDIs. Nonetheless, since many still unknown DDIs certainly exist and our analysis only covers DDIs known in 2011 (see section 4.4), the true importance of the DDI phenomenon is likely larger than what we observed. In addition, the relatively short study periods for Blumenau and Indianapolis compared to Catalonia may mask shifts in drug availability policy. This certainly highlights the importance of pursuing future studies with longer periods of observation as data becomes available.

In summary, our large-scale epidemiological analysis shows that DDIs are certainly a problem that affect a substantial proportion of patients in the three distinct populations studied. Ours is the first study to compare the DDI phenomenon in three large and distinct health care systems, both public and private, and follow close to 6 million patients for more than a decade. Because we studied very diverse populations and health systems, from developing to developed countries, our results likely generalize to a range of other nations where access to EHR data is still difficult or non-existent. Of particular importance is that similar gender and age biases exist in the administration of known DDIs in all observed public health systems, albeit with some context-specific differences we also characterize. Thus, physicians, drug developers, and health care professionals should be aware that the existence of sex and age biases need to be taken into consideration in drug management. The analysis, results, and tools we provide, can be used by others to investigate additional actionable interventions. Indeed, our study emphasizes that much more attention should be put to understand and reduce the DDI phenomenon and its biases. Because interactions between cultural, economical, and biological factors are likely at play, in addition to computational and epidemiological studies such as ours, the DDI phenomenon calls for greater interdisciplinary collaboration. We hope that by uncovering such a large footprint of the DDI phenomenon, with the burden it represents to patients and health care systems alike, we also contribute to awareness of the need to accelerate disruptive drug research toward new and safer therapeutic targets, particularly for chronic conditions.

## 4. Methods

### 4.1 Data – Blumenau

Drugs reported in the *Pronto* HIS are available via medical prescription only, free of charge, and administered to citizens of Blumenau. Via *Pronto*, doctors prescribe medications by selecting drugs and dosages, then pharmacists dispense them by selecting quantity. This allows us to estimate the length of drug administration in days. We note patients are not required to retrieve drugs from the public system. They can buy prescribed medications from private pharmacies at their own expense, without such transactions being recorded in *Pronto*. Drug names originally in Portuguese have been translated to English, disambiguated, and matched to their IDs in DrugBank, an open-source drug database that contains DDI information. Medications with multiple drug compounds have been split into their constituent drugs. Administered substances not matched in DrugBank were discarded. The data includes eighteen months (Jan 2014–Jun 2015) of anonymized drug administration and patient demographics retrieved from *Pronto*. It is the same data used in Correia *et al*. [5] except for the removal of ophthalmological drugs, topical drugs, and vaccines from the analysis. In total, we analyze 140 unique DrugBank IDs dispensed to 133,047 patients. The study was approved by Indiana University’s Institutional Review Board.

### 4.2 Data – Catalonia

Monthly drug billing data from the HIS includes drugs identified by their Anatomical Therapeutic Chemical (ATC) classification, which contains five levels of detail. We use the finest detail level—chemical substance—and remove topical drugs. For comparison, we map ATC codes to DrugBank IDs. Importantly, we note that: (a) a drug can map to more than one ATC code when it has different routes of administration or therapeutic uses, and (b) some ATC codes represent combined drugs. For simplicity, we aggregate all ATC code billing that matches a DrugBank ID and split combined drugs into their constituent drugs. Only patients born before January 2007 were included in the study. The data includes eleven years (Jan 2008-Dec 2018) of anonymized drug billing data, disease diagnoses (ICD-10), and patient demographics provided by the Catalan Health Institute and extracted from the Information System for the Development of Research in Primary Care. In total, we analyze 814 unique DrugBank IDs administered to 5,555,924 patients. The study was approved by the Jordi Gol University Institute for Research Primary Healthcare ethics committee.

### 4.3 Data – Indianapolis

Unlike the other populations, drugs in Indianapolis could have been administered as prescribed by primary physicians as well as administered in a hospital setting. The data includes drug quantity and treatment duration that allowed us to estimate the length of administration in days. Similarly to the Blumenau data, we disambiguate individual medication names and matched them to DrugBank IDs, and split medications with multiple drug compounds into their constituent drugs. We removed ophthalmological drugs, topical drugs, and vaccines from the analysis. The data includes two years (Jan 2017–Dec 2018) of anonymized drug administration data, disease diagnoses (ICD-10), and patient demographics from the Regenstrief Institute for all three care levels of a major health care provider in the city of Indianapolis. We analyze 1,228 unique DrugBank IDs dispensed to 264,607 patients. The study was approved by Indiana University’s Institutional Review Board.

### 4.4 Drug-drug interactions

To ensure all DDIs found from the earliest dispensation dates in our study to the most recent, we use the 2011 version of DrugBank as our drug interaction reference. Following the notation proposed in Correia *et al*. [5], we denote patients by *u* ∈*U*, and drugs by *i,j* ∈*D*, where *U*_*i*_ ∈*U* represents the subset of patients dispensed drug *i*, and *D*^*u*^ ⊆ *D* is the subset of drugs dispensed to patient *u*. Since patients can be administered a drug *i* multiple times during the study period, we denote the set of distinct administration intervals *a*^*i,u*^_*n*_ (in days or months) of drug *i* to patient *u* as 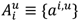. The total number of administrations and time units a patient *u* is administered a drug *i* are denoted by 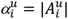 and 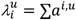. For Blumenau and Indianapolis we are able to compute drug administration length in days. For Catalonia, however, we only have monthly drug billing data, therefore, in this case, 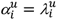 denotes the number of months drug *i* was administered to patient *u*. We assume dispensed drugs were administered for the entire prescribed length. Similarly, the number of distinct co-administration periods of two drugs (*i* and *j*) to patient *u* and the length of co-administration are denoted by 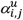 and 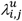, respectively (see fig. 5). To simply flag whether patient *u* co-administered drug pair (*i,j*) at least once, we define a Boolean variable 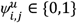:

**Figure 5.**
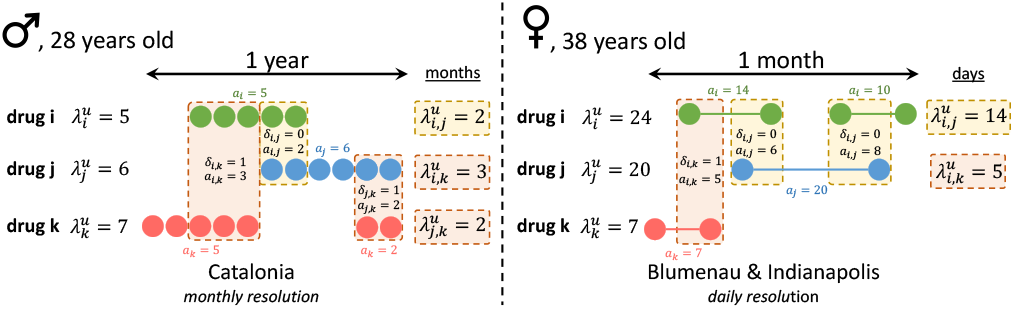
Diagram of co-administration and interaction computation for Catalonia, Blumenau, and Indianapolis. Two hypothetical patient-drug dispensing timelines with three drugs (*i, j*, & *k*) are represented. In Catalonia (left), two drugs (*i,j*) are assumed to be co-administered if they were dispensed and billed during the same month. In Blumenau and Indianapolis (right), two drugs are assumed to be co-administered if they were dispensed for an administration period with an overlap of at least one day. Drug administration lengths (in days for Blumenau and Indianapolis, and months for Catalonia) are shown for each dispensation. The three possible pairwise comparisons (*i,j*), (*i,k*), and (*j,k*) between the dispensed drugs are shown with their co-administration overlap marked with backgrounds in either orange (not known DDI) or red (known DDI).

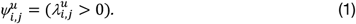

Likewise, to flag the co-administration to patient *u* of drugs (*i,j*) that are known to interact we define another Boolean variable 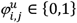:

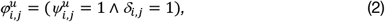

where the symmetrical binary map Δ: *DxD* → {0,1} indicates whether every drug pair (*i,j*) ∈*D* × *D* is a known DDI in DrugBank (*δ*_*i,j*_ = 1) or not (*δ*_*i,j*_ = 0).

For each observed DDI (∀*φ*_*i,j*_ = 1), we manually retrieve a severity score *s* ∈{*major, moderate, minor, n*/*a*} from drugs.com [16], a website containing drug information, including DDI descriptions, powered by the American Society of Health-System Pharmacists and IBM Watson Micromedex. From these values, we compute other quantities and sets per patient *u*, drug *i*, or drug pair (*i,j*).

To characterize the *conditional likelihood of a drug pair* (*i,j*) in the population, we obtain the number of patients who administered the drug pair concomitantly, 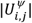, and normalize it by the number of patients who administered one of the drugs in the pair, to obtain the probability that patients who administered drug *i* also co-administered drug pair (*i,j*):

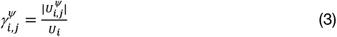

Values of 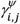 closer to 1 indicate that drug *j* is usually co-administered with drug *i* in the population, or vice-versa for 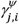, as this measure is not symmetrical 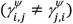.

Since 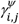 does not differentiate if drugs *i* and *j* are concomitantly administered for a short or long period of the time, and we assume that the length of DDI administration is relevant for ADR, we also characterize the length of co-administration of drug pairs to a patient *u* by calculating

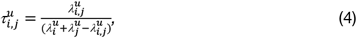

Where 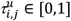. This measure of *normalized co-administration length* per patient differentiates between drug pairs with complete temporal overlap, 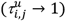, and with a small temporal overlap 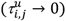. Its mean value for the cohort of patients who administered drug pair (*i,j*) concomitantly yields a measure of *strength of co-administration* of the pair in the population:

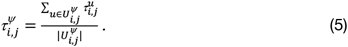

This proximity measure defines a weighted, undirected graph *T*^*ψ*^ [31] on set *D* with edges, 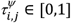, that relate drugs in the patient population according to the strength of co-administration (as inferred by normalized co-administration length). Graph *T*^*ψ*^ synthesizes the multivariate phenomenon of drug co-administration in a given population.

To focus on the DDI phenomenon, we compute the *strength of a DDI* as:

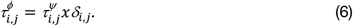

It also defines *T*^*ϕ*^, a subgraph of *T*^*ψ*^ that contains only the known interacting drug pairs observed in a given population (e.g., fig. 3). Graph *T*^*ϕ*^ thus synthesizes the multivariate DDI phenomenon in a given population as a network, which is further refined in section 4.7. Naturally, a *conditional likelihood of drug interaction* can be similarly obtained from *eq. 3* as

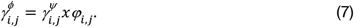

As noted above, this measure does not take into account length of DDI administration, while *eq. 6* does.

To test the significance of the observed DDIs in the population, we calculate Fisher’s exact tests on the number of patients affected by each DDI, 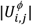, and the Bonferroni adjusted *p*−value based on the total number of DDI found in each population, 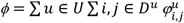. Interacting drug pairs with a false discovery rate (FDR) ≤ 0.05 are considered significant and further analyzed.

For each population we calculate the *risk of co-administration* (*RC*) as the number of patients who co-administered at least two drugs divided by the total number of patients, |*U*^*ψ*^|/|*U*|. Similarly, we calculate the *risk of interaction* (*RI*) as |*U*^*ϕ*^|/|*U*|, denoting the risk of any patient in the population to be administered at least one DDI.

### 4.5 Sex risks

The relative risk of co-administration (*RRC*) for women is computed as the ratio of the conditional probabilities of patients administering at least one pair of drugs concomitantly, given their sex:

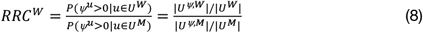

The same risk for men is computed as *RRC*^*M*^ = 1/*RRC*^*W*^. Similarly, we also compute the relative risk of interaction (*RRI*) for women as:

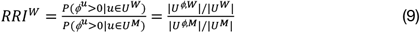

with *RRI*^*M*^ = 1/*RRI*^*W*^. Additionally, Fisher’s exact tests are used to calculate the significance of the various relative risk measures.

### 4.6 Age risks

To evaluate the effect of patient age in the DDI phenomenon, we bin patients into 5-year age groups (or age cohorts) to compute an age-dependent risk of co-administration and DDI. In other words, the risk of co-administration for age group [*y*1, *y*2] can be computed by simply constraining *RC* per age group:

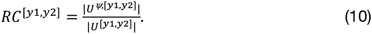

Similarly, the risk of interaction for age group [*y*1, *y*2] is calculated as

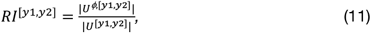

which we interpret as the risk of a patient in age range [*y*1, *y*2] being exposed to a known DDI. Additionally, we parse age risk by sex and drug pair, by computing *RI*^[*y*1,*y*2],*g*^ for each sex *g* ∈{*W, M*} using *eq. 11* for users *u* ∈*U*^[*y*1,*y*2],*g*^, and 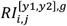 for each drug pair *i,j*. These allow us to also compute relative risks constrained on age ranges, sex, and drug pairs, such as *RRI*^[*y*1,*y*2],*W*^ and 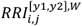. Note that due to the temporal nature of our study, patient age is calculated based on their date of birth and the date of the drug event. This means that individual patients may be accounted for in multiple independent age ranges.

### 4.7 Drug-drug interaction network

To synthesize, depict, and analyze the DDI phenomenon captured by the EHR data, we build a DDI network for each population where nodes represent drugs and edges denote an observed and significant (per criterion in section 4.4) drug interaction in the population. Each population network is defined by graph *T*^*ϕ*^, further refined such that edge width is proportional to *φ*_*i,j*_, the strength of DDI (*eq. 6*), while edge color represents the sex-specific risk (*eq. 9*, but computed for each DDI as 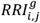) for women in darker red and men in darker blue. Further, node size denotes the probability of patients who administered drug *i* to be exposed to a DDI associated with that drug, and is computed as

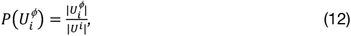

where 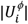 is the number of patients who administered a DDI involving drug *i*. An interactive application that allows the user to filter results and explore the associated network is available at http://disease-perception.bsc.es/ddinteract/.

### 4.8 Null model

The null model, 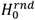 captures the expected increase in *RI*^[*y*1,*y*2]^ with age, given observed polypharmacy and patient demographics within each age group. We assume a random administration of drugs to patients in a specific age group, therefore maintaining the same number of unique drugs dispensed and co-administered for each randomly drawn patient. Specifically, we randomly draw |*U*^[*y*1,*y*2]^| patients from each age group [*y*1, *y*2]. Then for each patient *u*, we randomly “dispense” |*D*^*u*^| drugs drawn from set of drugs that were observed to be dispensed to patients in the same age group, *D*^[*y*1,*y*2]^. In other words, in the null model patients “administer” the same number of drugs as in the observed real population, but the drugs are randomly selected from the set of drugs observed to be prescribed for that age group. Then, *eq. 11* is calculated for the null model simulation and denoted by 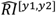. A risk disparity between the actual data and the null model can be computed as the relative risk

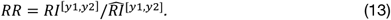

Additionally, Fisher’s exact tests are used to calculate the significance of the relative risk measures. Furthermore, the null model also uses the same number of “co-administred” 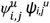 drug pairs (*i,j*) as observed in the real data, with the co-administered drugs *j* also drawn randomly from the set *D*^*u*^ of “administered” drugs to user *u* in the null model. These random drug pairs are subsequently checked for DDI status in DrugBank, as in the original analysis. We repeat this random sampling process 100 times and compute all derived risk measures, as done with the original data.

### 4.9 Removal of Omeprazole-associated interactions

Since Omeprazole is known to be over prescribed and has one of the largest number of interactions observed in our study (see tables S3 and S9), we simulate the replacement of Omeprazole with alternative PPIs in observed DDI cases. We use the ATC drug classification system that describes chemical subgroups containing drugs that could, in principle, be interchanged for the treatment of the same disease to identify alternatives. Thus, as proof of concept, we focus on the PPI subgroup: Omeprazole, Pantoprazole, Esomeprazole, Lansoprazole, and Rabeprazole. We then replace, in each situation, Omeprazole with the alternative that avoids interactions with other drugs and recalculate the previously described risk measures.

## Data Availability

No new data have been produced in this study; the data analyzed have been provided by different institutions.

http://disease-perception.bsc.es/ddinteract/

## Data Availability

Data is available at: https://github.com/rionbr/DDI-Cat-Indy-Bnu

## Author Contributions

J.S.V., R.B.C, L.M.R, and A.V conceived the research strategy. J.S.V and R.B.C conducted the analysis, and wrote the manuscript. M.C.A and R.L. helped interpret the results. R.B.C. and M.M.M acquired the Blumenau data. R.B.C and L.M.R acquired the Indianapolis data. J.S.V, R.L, and A.V. acquired the Catalonia data. All authors discussed the results. All authors approved the final manuscript.

## Declarations

## Acknowledgements

The authors would like to thank Deborah Rocha for copy editing, the city of Blumenau and the *Pronto* HIS for the Blumenau data, the Information System for the Development of Research in Primary Care for the Catalonia data, and the Regenstrief Institute for the Indianapolis data.

