## Supplementary Material for "Analysis of electronic health records from three distinct and large populations reveals high prevalence and biases in the co-administration of drugs known to interact"

### Supplemental Material

#### S1 Additional results on observed drug-drug interactions

To study the details of the DDI phenomenon in each population, here we present results using the entire temporal data: 18 months for Blumenau, 11 years for Catalonia, and 2 years for Indianapolis. We first observe that 211, 1,782, and 1,483 unique DDI were administered in the Blumenau, Catalonia, and Indianapolis populations, respectively. The top DDIs (and their severity) for each population are shown in table S3, and the entire list is available at http://disease-perception.bsc.es/ddinteract/.

The concomitant use of Omeprazole with various benzodiazepines (Clonazepam, Diazepam, and Alprazolam) is among the most frequent DDIs in Blumenau and Catalonia and is associated with higher risk for women in both cities. In Indianapolis, in contrast, the most frequent DDIs are the concomitant use of: two anticoagulants (Acetylsalicylic acid (ASA) Heparin), an anticoagulant and insulin (ASA combined with Insulin lispro or Insulin glargine), and a beta-blocker and a local anesthetic (Metoprolol and Lidocaine), all of which are associated with greater risk (*RI*) for men.

Drug availability differs in the three health care systems. Considering only the common set of 106 drugs dispensed in all three populations (fig. S4a), Catalonia is the population with the largest number of administered unique DDIs (206), followed by Indianapolis (183), and Blumenau (175), as shown in see fig. S4b. In addition, five DDIs are only observed in Catalonia—all associated with epilepsy treatment. Three of the interactions involve Phenytoin (co-administered with Disulfiram, Betamethasone and Mebendazole) and the two others involve Phenobarbital (co-administered with Betamethasone and Nifedipine). Interestingly, three of the four DDIs only observed in Blumenau and Indianapolis also involve epilepsy-treatment drugs in different combinations from Catalonia: Valproic acid (co-administered with Erythromycin, Phenytoin, and Phenobarbital).

Next, we select DDIs that are common to all three populations and highlight the drugs that are more frequently given with its interacting partner (see table S4), in regards to the number of patients they affect. As shown in table S4, Digoxin is the drug most often administered to patients in conjunction with its interacting drugs in Blumenau and Indianapolis (note that $\gamma_{i,j}^{\Phi} > \gamma_{j,i}^{\Phi}$, highlighted in bold in table S4). For instance, in the three populations, from all patients who administered Digoxin, 47-60% of them also co-administered Furosemide. Conversely, for all patients who administered Furosemide, only 4-12% also co-administered Digoxin. This DDI also has one of the largest observed strength of drug interaction ($\tau_{i,j}^{\Phi}$, table S4), which shows that it tends to be administered for long periods of time, increasing the risk of hospitalization due to Digoxin intoxication [32].

In addition, 10 out of the 12 shared DDIs (table S4) are related to cardiovascular disorders. Only two pairs make up the exception: Valproic acid–Carbamazepine and

Haloperidol–Lithium cation. The former are anticonvulsants usually prescribed to treat seizure and bipolar disorders and given in combination to boost mood stabilization when monotherapy using either drug fails [33]. The latter are anti-psychotic drugs used to treat schizophrenia and bipolar disorder, and combined to provide modest, statistically significant benefits in the treatment of schizoaffective disorder [34]. Even if both drugs are not frequently given together, our results denote a stronger association of Lithium cation with Haloperidol rather than the other way around, potentially due to the smaller effectiveness of Lithium alone compared with other neuroleptics.

Finally, we find that half of the shared DDIs pose a major health risks, such as hyperkalemia and kidney failure (Spironolactone-Losartan), increased risk of bleeding (Warfarin-Amiodarone), and excess mortality (DigoxinAmiodarone) [35].

#### S2 Strength of the co-administration

Table S1 shows the DDIs that are co-administered more than expected by change and are common in the three populations, ranked based on the strength of their association and the number of patients affected by them (see section 4.4). Half of these interactions pose a major risk for health, leading to hyperkalemia and kidney failure (Spironolactone and Losartan), increased risk of bleeding (Warfarin and Amiodarone), and excess mortality (Digoxin and Amiodarone) [35].

The highest ranked DDI pair is Digoxin (cardiac glycoside) with Furosemide (diuretic) combination, which significantly increases the risk of hospitalization for Digoxin intoxication [32]. In addition to Furosemide, Digoxin is significantly associated also with two drugs used to treat high blood pressure associated to heart failure (Spironolactone and Carvedilol), an antiarrhythmic drug (Amiodarone), and Levothyroxine, a drug used to treat thyroid hormone deficiency. It has been shown that, even at subclinical levels, hypothyroidism worsens heart failure prognosis [36].

**Table S1.** The 12 drug-drug interactions administered more than expected and common to the Blumenau, Indianapolis, and Catalonia populations, ordered based on the rank product of the strength of the association ($\tau_{i,j}$) and the number of patients (%) in the three populations. To allow cross population comparison, number of patients are shown as a percentage of the overall patient population ($|U^{\phi}_{i,j}|/|U|$).

| $i$ | $j$ | $\tau^{\text{Bnu}}_{i,j}$ | $\tau^{\text{Cat}}_{i,j}$ | $\tau^{\text{Indy}}_{i,j}$ | % Bnu | % Cat | % Indy | severity |
|---|---|---|---|---|---|---|---|---|
| Digoxin | Furosemide | 0.601 | 0.271 | 0.835 | 0.288 | 0.656 | 0.168 | Moderate |
| Spironolactone | Losartan | 0.623 | 0.256 | 0.771 | 0.500 | 0.108 | 0.174 | Major |
| Diltiazem | Amiodarone | 0.630 | 0.275 | 0.882 | 0.019 | 0.042 | 0.087 | Major |
| Digoxin | Spironolactone | 0.578 | 0.294 | 0.849 | 0.204 | 0.184 | 0.055 | Minor |
| Spironolactone | Captopril | 0.512 | 0.307 | 0.987 | 0.040 | 0.016 | 0.012 | Major |
| Digoxin | Carvedilol | 0.517 | 0.258 | 0.854 | 0.130 | 0.110 | 0.061 | Moderate |
| Digoxin | Amiodarone | 0.564 | 0.260 | 0.859 | 0.039 | 0.082 | 0.064 | Major |
| Valproic acid | Carbamazepine | 0.410 | 0.375 | 0.777 | 0.115 | 0.029 | 0.006 | Moderate |
| Warfarin | Amiodarone | 0.591 | 0.260 | 0.847 | 0.069 | 0.040 | 0.083 | Major |
| Digoxin | Levothyroxine | 0.584 | 0.253 | 0.851 | 0.077 | 0.079 | 0.057 | Moderate |
| Allopurinol | Warfarin | 0.510 | 0.207 | 0.911 | 0.035 | 0.023 | 0.054 | Moderate |
| Haloperidol | Lithium cation | 0.462 | 0.263 | 0.849 | 0.111 | 0.010 | 0.009 | Major |

#### S3 Drug-drug interaction networks

Both Blumenau and Catalonia have larger number of distinct DDIs being co-administered to a higher proportion of women (see number of red edges in networks, such as fig. 3). In Blumenau, out of the 211 DDI observed 142 (67%) have an increased risk for women while only 69 (33%) have an increased risk for men. For Catalonia these numbers are 981 (55%) and 801 (45%), for women and men, respectively. For Indianapolis, even though the populationlevel risk of DDI is higher for women (as seen in table 1), there is almost the same proportion of distinct DDIs associated with an increased risk for both sexes, 729 (49.8%) for women and 744 (50.2%) for men. Interestingly, when only DDIs co-administered more than expected by chance are considered (Fisher's exact test, FDR ≤ 0.05), the proportions of sex-based relative risk DDIs are reversed in Blumenau and Catalonia. In Blumenau, out of the 48 significant DDI observed, 23 (48%) have an increased risk for women and 25 (52%) have an increased risk for men, a 67 − 48 = 19% decrease for women. For Catalonia these same numbers are 103 (42%) and 143 (58%), for women and men, respectively, a 13% decrease for women. And lastly for Indianapolis the proportions are 83 (38.6%) significant DDIs for women and 132 (61.4%) for men, also a decrease of 11.1% to women. Naturally, a significant DDI proportion reduction for women denote an increase for men.

**Table S2.** Relative risk for women ($RRW$) of drug co-administration ($RRC^W$) and interactions ($RRI^W$); the latter is also computed for types of interactions as per drugs.com (minor, moderate, and major). The percentage of patients of each sex (M = man; W= woman) for each case is also shown. Values shown for all three populations for the whole study period, that is, 18 month for Blumenau, 11 years for Catalonia, and 2 years for Indianapolis.

| | Blumenau | | Catalonia | | Indianapolis | |
|---|---|---|---|---|---|---|
| | RRW | % W - M | RRW | % W- M | RRW | % W - M |
| Co-administration ($RRC^W$) | 1.07 | (79.08% - 74.06%) | 1.03 | (91.03% - 88.26%) | 1.06 | (77.33% - 73.13%) |
| Interaction ($RRI^W$) | 1.54 | (14.64% - 9.49%) | 1.35 | (23.3% - 17.21%) | 1.14 | (13.75% - 12.1%) |
| Minor Interaction | 0.81 | (0.36% - 0.44%) | 1.35 | (0.56% - 0.41%) | 1.04 | (1.23% - 1.18%) |
| Moderate interaction | 1.59 | (11.3% - 7.1%) | 1.53 | (18.85% - 12.31%) | 1.16 | (11.26% - 9.71%) |
| Major interaction | 1.53 | (6.25% - 4.07%) | 1.11 | (7.76% - 6.98%) | 1.04 | (5.32% - 5.13%) |

**Table S3.** Top 10 DDI observed in the three populations ranked based on the number of patients ($|U^{\phi}_{i,j}|$) taking them. $\tau^{\phi}_{i,j}$ denotes the strength of interaction (*eq. 6*). $RRI^W$ denotes the relative risk of interaction for women (*eq. 9*), with values larger than (smaller than) 1 denoting higher risk for women (men). The false discovery rate (FDR) and odds ratio represent the significance of the interaction, i.e. whether both drugs are administered more than expected by chance. DDI severity shown as retrieved from Drugs.com [16].

| Pop. | $i$ | $j$ | $\lvert U^{\Phi}_{i,j}\rvert$ | $\tau^{\Phi}_{i,j}$ | $RRI^W$ | odds | FDR | severity |
|---|---|---|---|---|---|---|---|---|
| Blumenau | Omeprazole | Clonazepam | 5,076 | 0.333 | 2.28 | 2.44 | 0 | Moderate |
| | Acetylsalicylic acid | Ibuprofen | 2,115 | 0.408 | 1.42 | 0.50 | 1 | Major |
| | Atenolol | Ibuprofen | 1,459 | 0.406 | 1.88 | 0.58 | 1 | Moderate |
| | Acetylsalicylic acid | Glyburide | 1,249 | 0.673 | 0.89 | 11.40 | 0 | Moderate |
| | Amitriptyline | Fluoxetine | 1,190 | 0.595 | 3.55 | 2.17 | 0 | Major |
| | Omeprazole | Diazepam | 998 | 0.346 | 1.21 | 1.79 | 0 | Moderate |
| | Fluconazole | Simvastatin | 892 | 0.328 | 2.64 | 0.83 | 1 | Major |
| | Spironolactone | Enalapril | 714 | 0.601 | 0.67 | 6.60 | 0 | Major |
| | Spironolactone | Losartan | 667 | 0.623 | 1.15 | 7.81 | 0 | Major |
| | Fluconazole | Clonazepam | 627 | 0.269 | 3.41 | 0.88 | 1 | None |
| Catalonia | Omeprazole | Diazepam | 253,473 | 0.223 | 1.66 | 0.31 | 1 | Moderate |
| | Omeprazole | Alprazolam | 167,507 | 0.201 | 2.13 | 0.42 | 1 | Moderate |
| | Acetylsalicylic acid | Ibuprofen | 133,691 | 0.092 | 0.88 | 0.11 | 1 | Major |
| | Acetaminophen | Acenocoumarol | 98,427 | 0.178 | 1.08 | 0.50 | 1 | None |
| | Prednisone | Acetylsalicylic acid | 49,297 | 0.116 | 0.77 | 0.47 | 1 | Moderate |
| | Omeprazole | Clonazepam | 47,811 | 0.183 | 1.51 | 0.65 | 1 | Moderate |
| | Furosemide | Ibuprofen | 44,911 | 0.138 | 1.52 | 0.07 | 1 | Moderate |
| | Bisoprolol | Ibuprofen | 44,276 | 0.083 | 1.00 | 0.09 | 1 | Moderate |
| | Alendronic acid | Ibuprofen | 43,869 | 0.113 | 11.40 | 0.17 | 1 | Moderate |
| | Acetylsalicylic acid | Gliclazide | 36,543 | 0.257 | 0.61 | 2.69 | 0 | None |
| Indianapolis | Acetylsalicylic acid | Heparin | 5,685 | 0.942 | 0.74 | 7.31 | 0 | Moderate |
| | Metoprolol | Lidocaine | 3,938 | 0.766 | 0.79 | 1.26 | 0 | Moderate |
| | Insulin lispro | Acetylsalicylic acid | 3,853 | 0.92 | 0.79 | 6.71 | 0 | NA |
| | Dexamethasone | Neostigmine | 3,438 | 0.969 | 1.40 | 17.10 | 0 | Moderate |
| | Insulin glargine | Acetylsalicylic acid | 2,463 | 0.855 | 0.76 | 4.73 | 0 | NA |
| | Metoprolol | Salbutamol | 2,358 | 0.776 | 1.01 | 0.92 | 1 | Moderate |
| | Prednisone | Acetylsalicylic acid | 2,240 | 0.739 | 1.08 | 1.03 | 1 | Moderate |
| | Acetylsalicylic acid | Dexamethasone | 2,019 | 0.541 | 1.22 | 0.62 | 1 | Moderate |
| | Ketorolac | Acetylsalicylic acid | 1,927 | 0.623 | 1.03 | 0.53 | 1 | Major |
| | Verapamil | Midazolam | 1,793 | 0.989 | 0.56 | 19.90 | 0 | Moderate |

**Table S4.** Drug-drug interactions co-administered significantly more than expected (see section 4.4) were 246 out of 1,782 in Catalonia, 5 out of 1,483 in Indianapolis, and 48 out of 211 in Blumenau. The 12 shown in the table are those common to the three populations, ranked by population "footprint" ($\gamma^{\phi}_{i,j}$ and $\gamma^{\phi}_{j,i}$ values). Values in bold denote the higher value between drugs *i* and *j*. Strength of drug interaction (*eq. 6*) is shown for each population. To allow cross population comparison, the number of patients is shown as a percentage of the overall patient population ($|U^{\phi}_{i,j}|/|U|$).

| $i$ | $i$ | Blumenau $\gamma^{\Phi}_{i,j}$ | $\gamma^{\Phi}_{j,i}$ | % | $\tau^{\Phi}_{i,j}$ | Catalonia $\gamma^{\Phi}_{i,j}$ | $\gamma^{\Phi}_{j,i}$ | % | $\tau^{\Phi}_{i,j}$ | Indianapolis $\gamma^{\Phi}_{i,j}$ | $\gamma^{\Phi}_{j,i}$ | % | $\tau^{\Phi}_{i,j}$ | severity |
|---|---|---|---|---|---|---|---|---|---|---|---|---|---|---|
| Digoxin | Furosemide | **0.60** | 0.12 | 0.29 | 0.6 | **0.46** | 0.09 | 0.66 | 0.27 | **0.49** | 0.04 | 0.17 | 0.84 | Moderate |
| Digoxin | Spironolactone | **0.42** | 0.14 | 0.2 | 0.58 | **0.13** | 0.12 | 0.18 | 0.29 | **0.16** | 0.03 | 0.06 | 0.85 | Minor |
| Digoxin | Carvedilol | **0.27** | 0.15 | 0.13 | 0.52 | 0.08 | **0.09** | 0.11 | 0.26 | **0.18** | 0.03 | 0.06 | 0.85 | Moderate |
| Digoxin | Amiodarone | **0.08** | 0.06 | 0.04 | 0.56 | 0.06 | **0.07** | 0.08 | 0.26 | **0.19** | 0.11 | 0.06 | 0.86 | Major |
| Warfarin | Amiodarone | 0.10 | **0.11** | 0.07 | 0.59 | **0.12** | 0.04 | 0.04 | 0.26 | 0.08 | **0.14** | 0.08 | 0.85 | Major |
| Haloperidol | Lithium cation | 0.14 | **0.16** | 0.11 | 0.46 | 0.01 | **0.04** | 0.01 | 0.26 | 0.01 | **0.06** | 0.01 | 0.85 | Major |
| Spironolactone | Losartan | **0.35** | 0.07 | 0.50 | 0.62 | **0.07** | 0.02 | 0.11 | 0.26 | **0.10** | 0.04 | 0.17 | 0.77 | Major |
| Diltiazem | Amiodarone | **0.03** | 0.03 | 0.02 | 0.63 | 0.04 | **0.04** | 0.04 | 0.28 | 0.08 | **0.15** | 0.09 | 0.88 | Major |
| Spironolactone | Captopril | **0.03** | 0.03 | 0.04 | 0.51 | 0.01 | **0.02** | 0.02 | 0.31 | 0.01 | **0.20** | 0.01 | 0.99 | Major |
| Valproic acid | Carbamazepine | **0.13** | 0.07 | 0.12 | 0.41 | 0.04 | **0.06** | 0.03 | 0.38 | 0.01 | **0.03** | 0.01 | 0.78 | Moderate |
| Digoxin | Levothyroxine | **0.16** | 0.01 | 0.08 | 0.58 | **0.06** | 0.02 | 0.08 | 0.25 | **0.17** | 0.01 | 0.06 | 0.85 | Moderate |
| Allopurinol | Warfarin | 0.04 | **0.05** | 0.04 | 0.51 | 0.01 | **0.07** | 0.02 | 0.21 | 0.05 | **0.05** | 0.05 | 0.91 | Moderate |

**Table S5.** Major DDI co-administered in the three populations with discordant sex-associated risk (interactions that are associated with a higher risk for women in one or two populations and a higher risk for men in the other). For instance, column "(Cat = Indy) ≠ Bnu" list major DDI with shared sex-associated risk in Catalonia and Indianapolis but reversed sex risk for Blumenau. Similarly, the same logic describes the other columns.

| (Cat = Indy) ≠ Bnu | (Cat = Bnu) ≠ Indy | (Indy = Bnu) ≠ Cat |
|---|---|---|
| Acetylsalicylic acid - Ibuprofen-man | Diclofenac - Warfarin-man | Diltiazem - Carbamazepine-men |
| Atenolol - Diltiazem-man | Fluoxetine - Lithium cation-woman | Diltiazem - Simvastatin-women |
| Atenolol - Verapamil-woman | Hydrochlorothiazide - Lithium cation-woman | Warfarin - Phenobarbital-men |
| Carbamazepine - Verapamil-woman | Spironolactone - Captopril-women | |
| Ciprofloxacin - Warfarin-man | Spironolactone - Enalapril-men | |
| Diltiazem - Amiodarone-man | Warfarin - Ibuprofen-men | |
| Fluconazole - Warfarin-women | | |
| Furosemide - Gentamicin-women | | |
| Haloperidol - Lithium cation-men | | |
| Propranolol - Verapamil-women | | |
| Warfarin - Metronidazole-men | | |

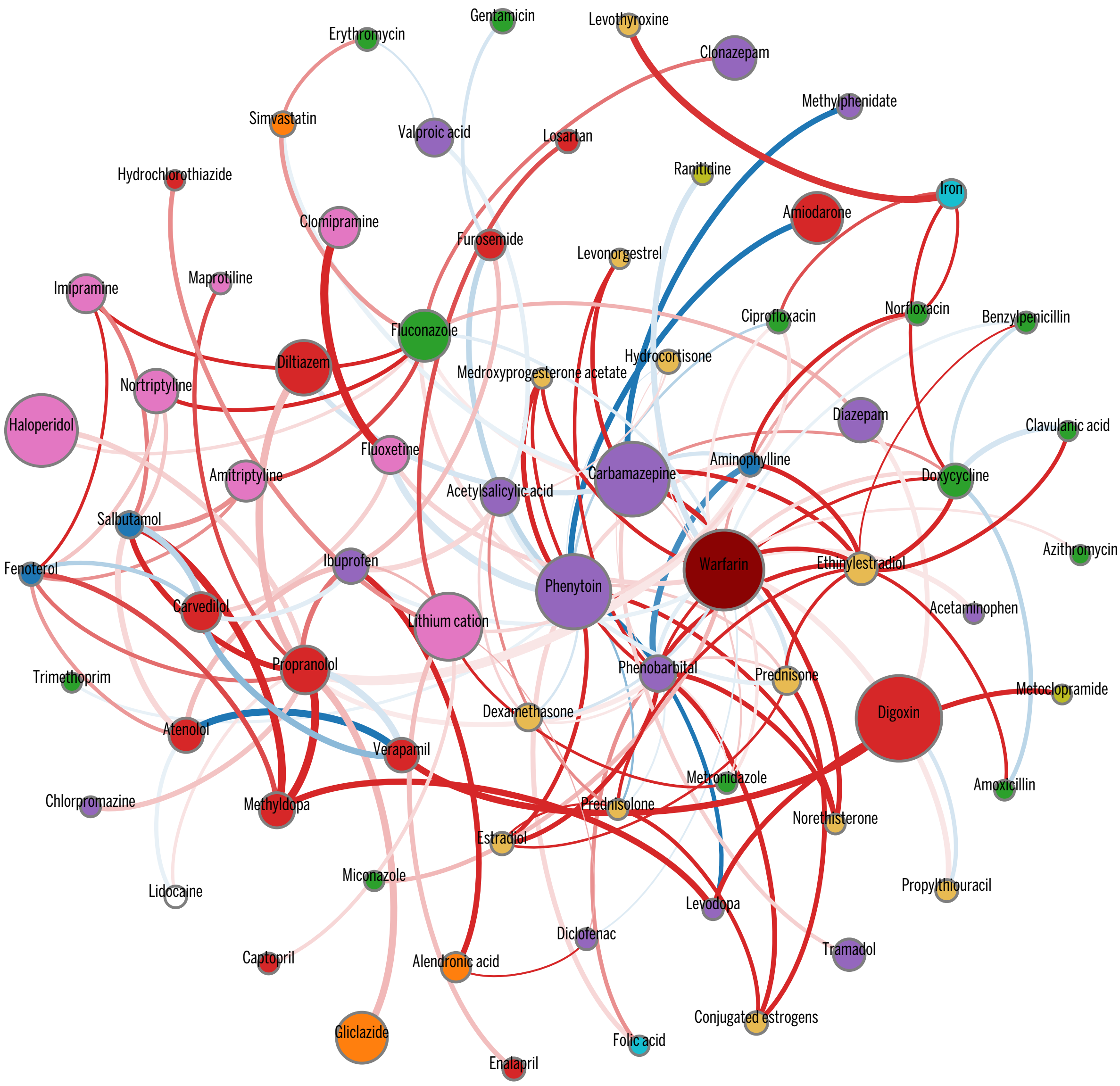


**Figure S1.** Blumenau DDI Network. A weighted version of network Δ where weights are defined by $\tau^{\phi}_{i,j}$. Nodes denote drugs $i$ involved in at least one co-administration known to be a DDI. Node color represents the highest level of primary action class, as retrieved from Drugs.com (see legend in fig. 3). Node size represents the probability of patients to be affected by a DDI involving the drug $P(U^{\phi}_{i})$ obtained from *eq. 12*. Edge weights are the values of $\tau^{\phi}_{i,j}$ obtained from *eq. 5*. Edge colors denote $RRI^{g}_{i,j}$, where $g \in M, W$, to identify DDI edges that are higher risk for women (red) or men (blue). Color intensity for $RRI^{g}_{i,j}$ varies in [1,5]; that is, values are clipped at 5.

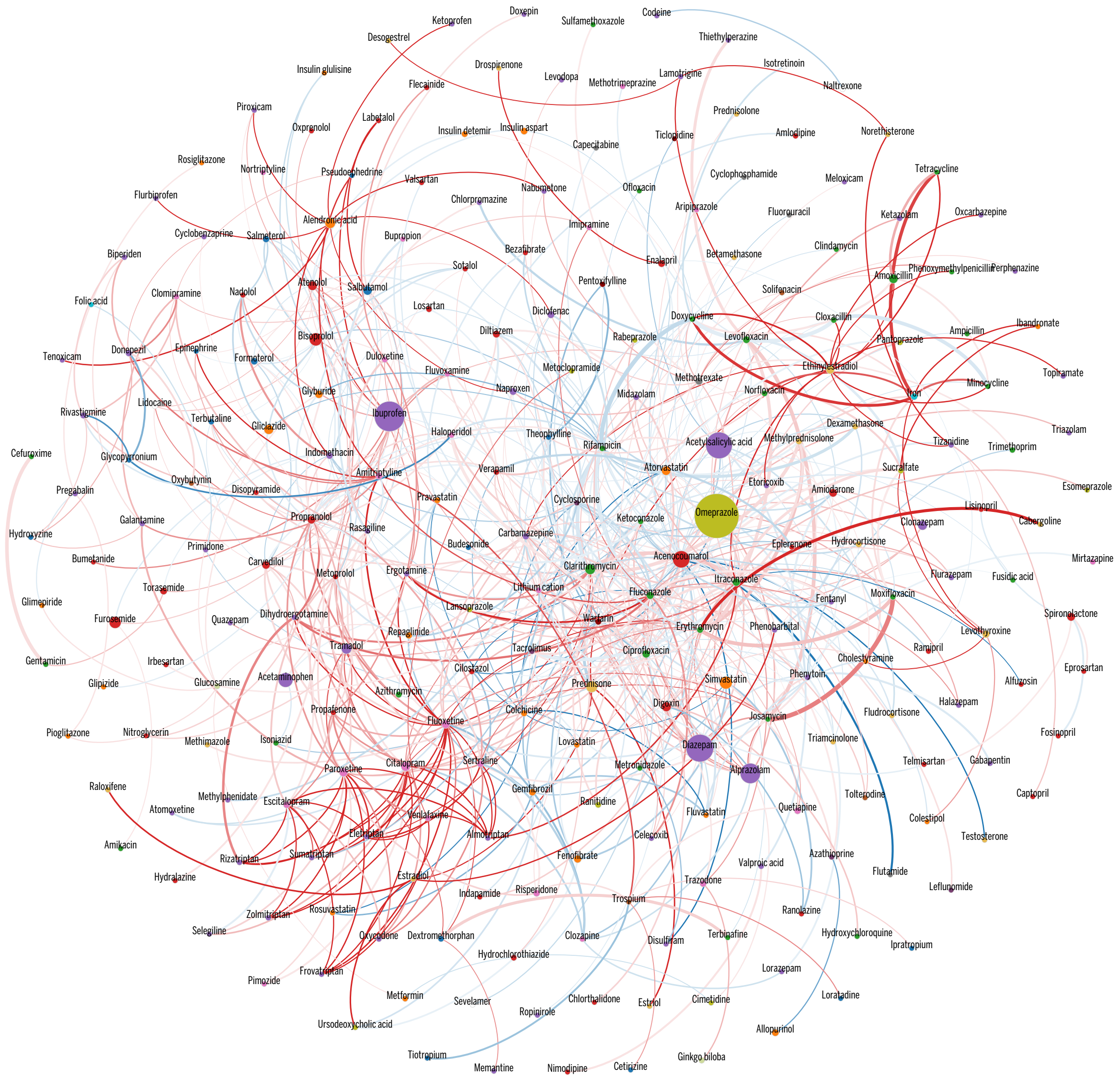


**Figure S2.** Catalonia DDI Network. A weighted version of network $\Delta$ where weights are defined by $\tau^{\phi}_{i,j}$. Nodes denote drugs *i* involved in at least one co-administration known to be a DDI. Node color represents the highest level of primary action class, as retrieved from Drugs.com (see legend in fig. 3). Node size represents the probability of patients to be affected by a DDI involving the drug $P(U^{\phi}_{i})$ obtained from *eq. 12*. Edge weights are the values of $\tau^{\phi}_{i,j}$ obtained from *eq. 5*. Edge colors denote $RRI^{g}_{i,j}$, where $g \in M, W$, to identify DDI edges that are higher risk for women (red) or men (blue). Color intensity for $RRI^{g}_{i,j}$ varies in [1,5]; that is, values are clipped at 5.

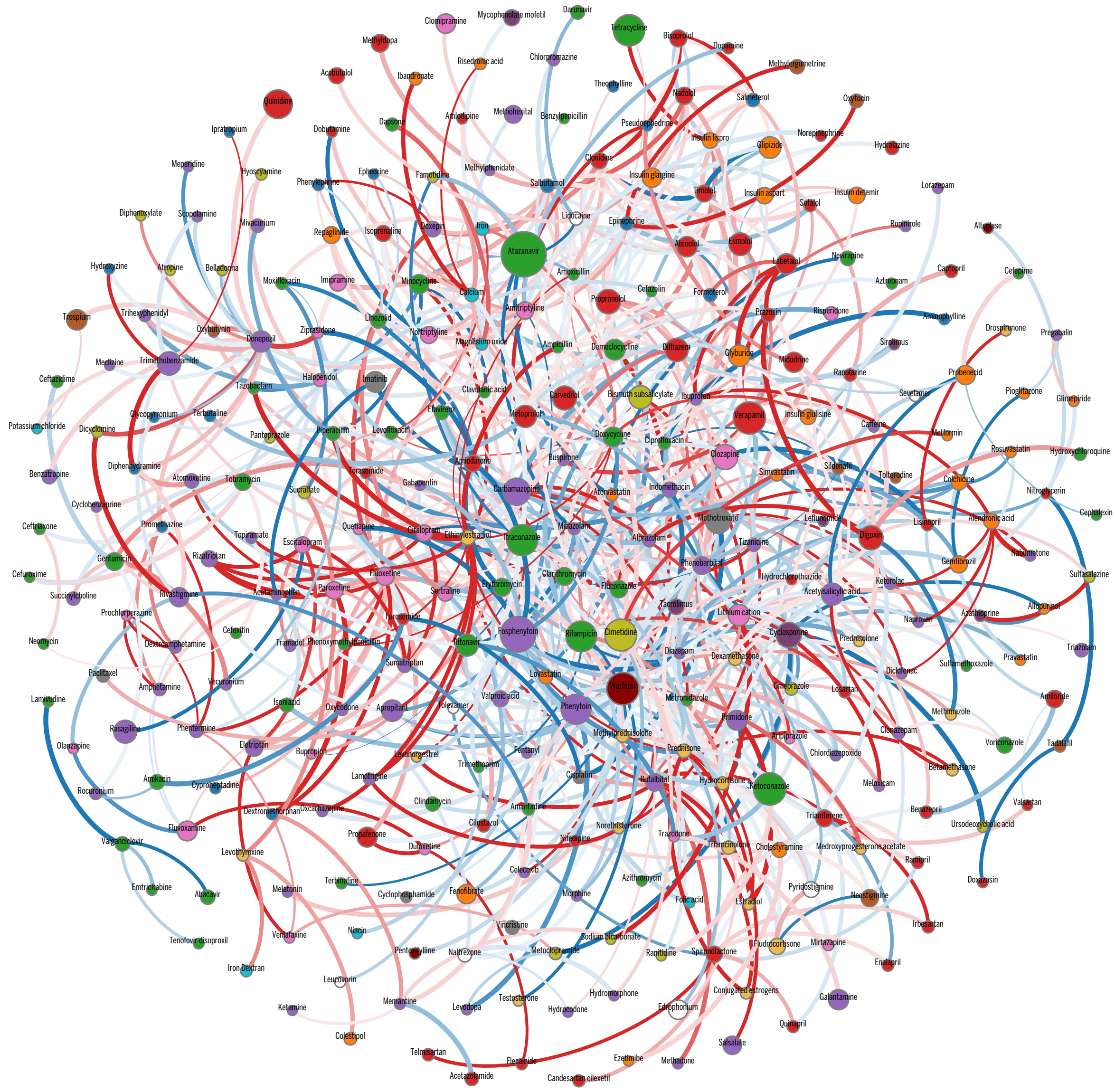


**Figure S3.** Indianapolis DDI Network. A weighted version of network Δ where weights are defined by $\tau^{\phi}_{i,j}$. Nodes denote drugs $i$ involved in at least one co-administration known to be a DDI. Node color represents the highest level of primary action class, as retrieved from Drugs.com (see legend in fig. 3). Node size represents the probability of patients to be affected by a DDI involving the drug $P(U^{\phi}_{i})$ obtained from *eq. 12*. Edge weights are the values of $\tau^{\phi}_{i,j}$ obtained from *eq. 5*. Edge colors denote $RRI^{g}_{i,j}$, where $g \in M, W$, to identify DDI edges that are higher risk for women (red) or men (blue). Color intensity for $RRI^{g}_{i,j}$ varies in [1,5]; that is, values are clipped at 5.

**Table S6.** Fisher's exact test results analyzing the significance of the association between omeprazole and diazepam/clonazepam compared to all the other proton pump inhibitors in Catalonia and Indianapolis. Arrows ($\leftarrow$ and $\rightarrow$) denote the direction of the odds-ratio and the p-values, where arrows pointing to the right denote a higher risk for omeprazole, while arrows pointing to the left denote a higher risk for the alternative proton pump inhibitors.

| | Benzodiazepine | $i$ | | substitute | odds-ratio | $p$-value |
|---|---|---|---|---|---|---|
| Catalonia | Diazepam | Omeprazole | $\rightarrow$ | together | 17,656 | 0 |
| | | | $\rightarrow$ | Pantoprazole | 16,039 | 0 |
| | | | $\rightarrow$ | Esomeprazole | 18,464 | 0 |
| | | | $\rightarrow$ | Rabeprazole | 19,138 | 0 |
| | | | $\rightarrow$ | Lansoprazole | 19,198 | 0 |
| | Clonazepam | Omeprazole | $\rightarrow$ | together | 12,228 | 9.71e-74 |
| | | | $\rightarrow$ | Pantoprazole | 10,967 | 3.93e-09 |
| | | | $\rightarrow$ | Esomeprazole | 13,356 | 2.95e-35 |
| | | | $\rightarrow$ | Rabeprazole | 12,859 | 1.62e-22 |
| | | | $\rightarrow$ | Lansoprazole | 13,438 | 4.14e-42 |
| Indianapolis | Diazepam | Omeprazole | $\leftarrow$ | together | 38,345 | 2.08e-62 |
| | | | $\leftarrow$ | Pantoprazole | 42,095 | 1.14e-68 |
| | | | $\leftarrow$ | Esomeprazole | 15,811 | 0.00774 |
| | | | $\leftarrow$ | Lansoprazole | 37,599 | 3.91e-19 |
| | Clonazepam | Omeprazole | $\leftarrow$ | together | 13,591 | 2.11e-05 |
| | | | $\leftarrow$ | Pantoprazole | 14,109 | 4.92e-06 |
| | | | $\leftarrow$ | Esomeprazole | 11,401 | 0.20403 |
| | | | $\leftarrow$ | Lansoprazole | 12,606 | 0.0698 |

**Table S7.** Relative risk of interaction for $(RRI_{i,j}^{[y1,y2],W})$, and risk of interaction for both women $(RI_{i,j}^{[y1,y2],W})$ and man $(RI_{i,j}^{[y1,y2],M})$ for the DDI between Alendronic acid and Ibuprofen by age range and sex (as defined in sections 4.5 and 4.6).

| | $RRI_{i,j}^{[y1,y2],W}$ | | | Blumenau | | Catalonia | | Indianapolis | |
|---|---|---|---|---|---|---|---|---|---|
| Age | Blumenau | Catalonia | Indianapolis | $RI_{i,j}^{[y1,y2],W}$ | $RI_{i,j}^{[y1,y2],M}$ | $RI_{i,j}^{[y1,y2],W}$ | $RI_{i,j}^{[y1,y2],M}$ | $RI_{i,j}^{[y1,y2],W}$ | $RI_{i,j}^{[y1,y2],M}$ |
| 00-04 | 0 | Inf | 0 | 0 | 0 | 0.004 | 0 | 0 | 0 |
| 05-09 | 0 | 0 | 0 | 0 | 0 | 0 | 0.001 | 0 | 0 |
| 10-14 | 0 | 0.53 | Inf | 0 | 0 | 0.002 | 0.004 | 0.012 | 0 |
| 15-19 | 0 | 2.72 | 0 | 0 | 0 | 0.007 | 0.002 | 0 | 0 |
| 20-24 | 0 | 1.67 | 0 | 0 | 0 | 0.005 | 0.003 | 0 | 0 |
| 25-29 | 0 | 1.42 | 0 | 0 | 0 | 0.005 | 0.004 | 0 | 0 |
| 30-34 | Inf | 2.42 | 0 | 0.034 | 0 | 0.012 | 0.005 | 0 | 0 |
| 35-39 | 0 | 1.93 | 0 | 0 | 0 | 0.016 | 0.008 | 0 | 0 |
| 40-44 | 1.22 | 2.96 | 0 | 0.062 | 0.051 | 0.038 | 0.013 | 0 | 0 |
| 45-49 | Inf | 5.31 | 0 | 0.25 | 0 | 0.126 | 0.024 | 0 | 0.014 |
| 50-54 | 17.25 | 12.76 | 0 | 0.382 | 0.022 | 0.477 | 0.037 | 0 | 0 |
| 55-59 | 33.08 | 19.07 | Inf | 0.816 | 0.025 | 1.128 | 0.059 | 0.009 | 0 |
| 60-64 | 6.15 | 17.95 | Inf | 1.341 | 0.218 | 1.81 | 0.101 | 0.037 | 0 |
| 65-69 | 9.16 | 15.13 | 5.22 | 1.989 | 0.217 | 2.143 | 0.142 | 0.078 | 0.015 |
| 70-74 | 6.92 | 11.31 | Inf | 2.537 | 0.367 | 2.008 | 0.178 | 0.048 | 0 |
| 75-79 | 5.92 | 9.14 | Inf | 2.108 | 0.356 | 1.893 | 0.207 | 0.022 | 0 |
| 80-84 | 6.21 | 6.72 | Inf | 1.699 | 0.274 | 1.544 | 0.23 | 0.06 | 0 |
| 85-89 | 0.88 | 5.32 | Inf | 0.241 | 0.273 | 0.998 | 0.187 | 0.044 | 0 |
| 90+ | Inf | 4.15 | 0 | 1.183 | 0 | 0.689 | 0.166 | 0 | 0 |

**Table S8.** Number and percentage of patients who administered Alendronic acid, Ibandronate, Risedronatem, and Zoledronic acid in Blumenau, Catalonia, and Indianapolis during the entire study period.

| | Number of patients | | | Percentage of patients | | |
|---|---|---|---|---|---|---|
| Bisphosphonates | Blumenau | Catalonia | Indianapolis | Blumenau | Catalonia | Indianapolis |
| Alendronic acid | 2,228 | 193,531 | 1,353 | 1.669 | 3.483 | 0.5 |
| Ibandronate | 0 | 33,295 | 91 | 0 | 0.599 | 0.034 |
| Risedronatem | 0 | 72,712 | 70 | 0 | 1.309 | 0.026 |
| Zoledronic acid | 0 | 0 | 269 | 0 | 0 | 0.099 |

**Table S9.** Number and percentage of patients who administered Omeprazole in Blumenau, Catalonia, and Indianapolis.

| | Number of patients | Percentage of patients |
|---|---|---|
| Blumenau | 24,495 | 18.35 |
| Catalonia | 2,640,806 | 47.53 |
| Indianapolis | 12,119 | 4.48 |

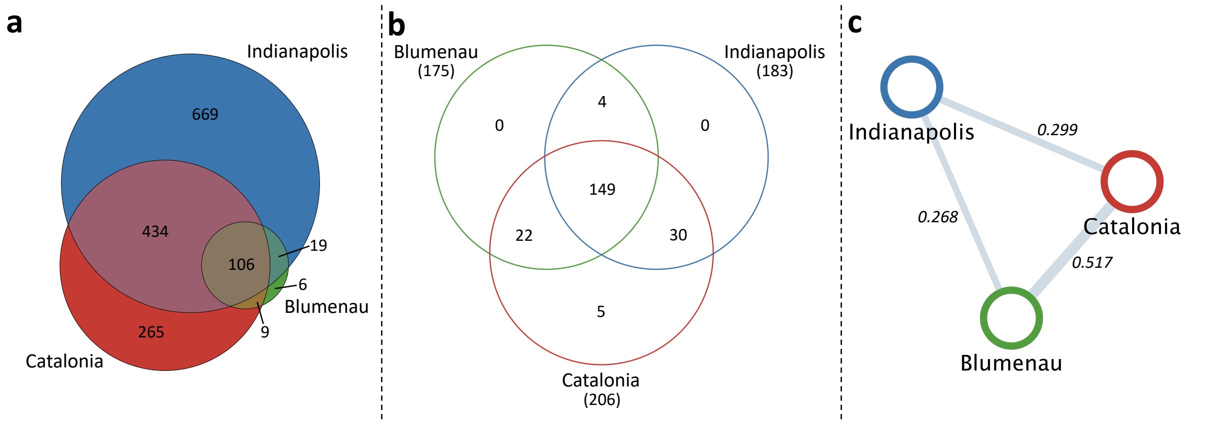


**Figure S4.** Numbers of drugs administered and drug-drug interactions in Blumenau, Catalonia, and Indianapolis. (a) Venn diagram representing the number of unique drugs dispensed in the three populations. (b) Venn diagram representing the number of unique drug-drug interactions in Blumenau, Catalonia and Indianapolis, when considering exclusively the 106 drugs administered in the three populations. (c) Spearman's correlations of the strength of interaction , $\tau_{i,j}^{\phi}$ (*eq. 6*), for all 149 DDI pairs observed in the shared set.

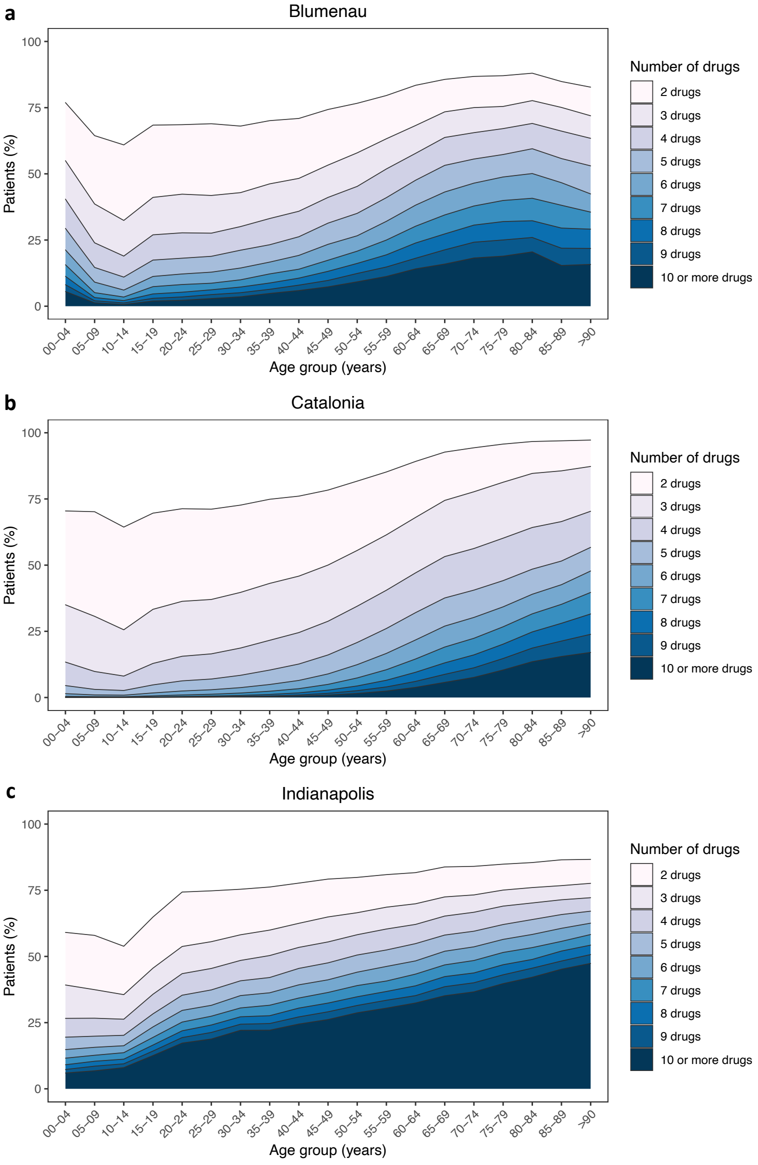


**Figure S5.** Percentage of patients co-administered 2 or more drugs simultaneously in Blumenau, Catalonia and Indianapolis.

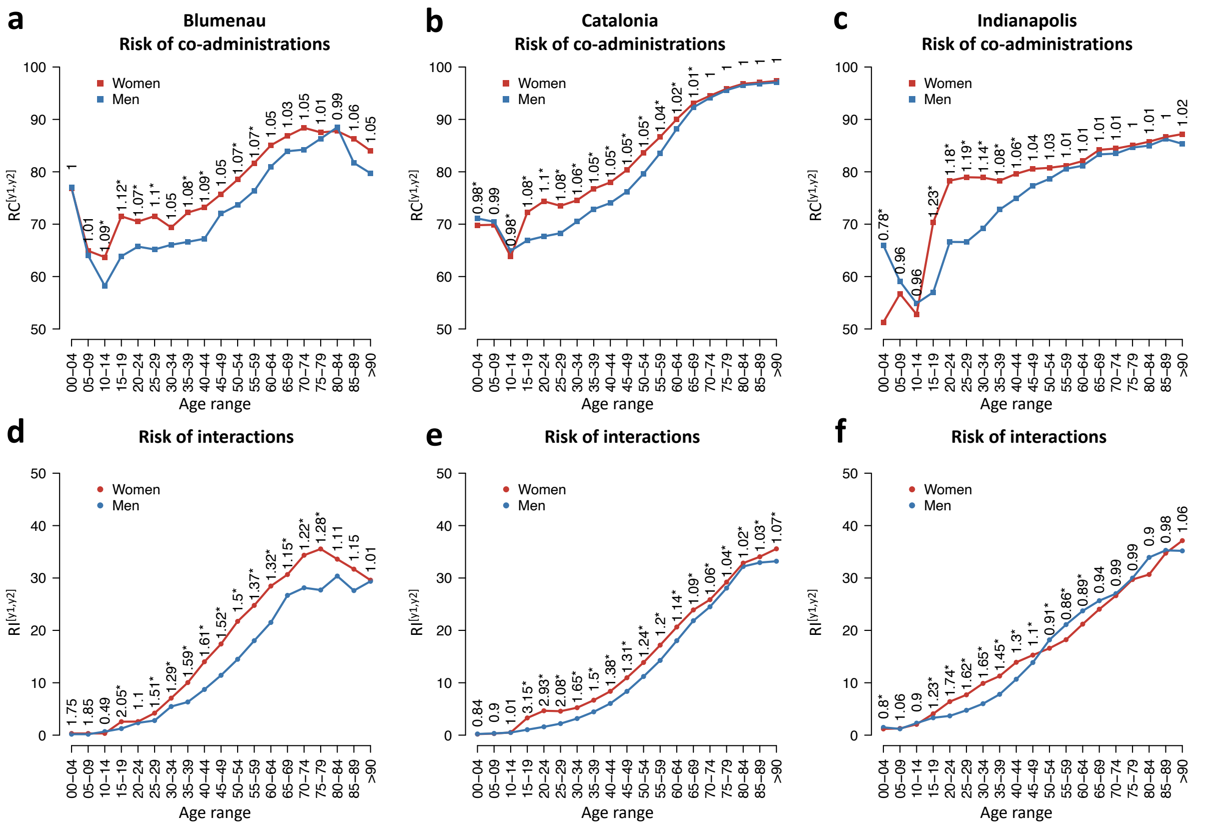


**Figure S6.** Risk of co-administration of drugs (a-c) and drugs known to interact (d-f) by age and sex in Blumenau, Catalonia, and Indianapolis analyzing the whole study period. Red and blue colors denote the risk of co-administration in women and men respectively.

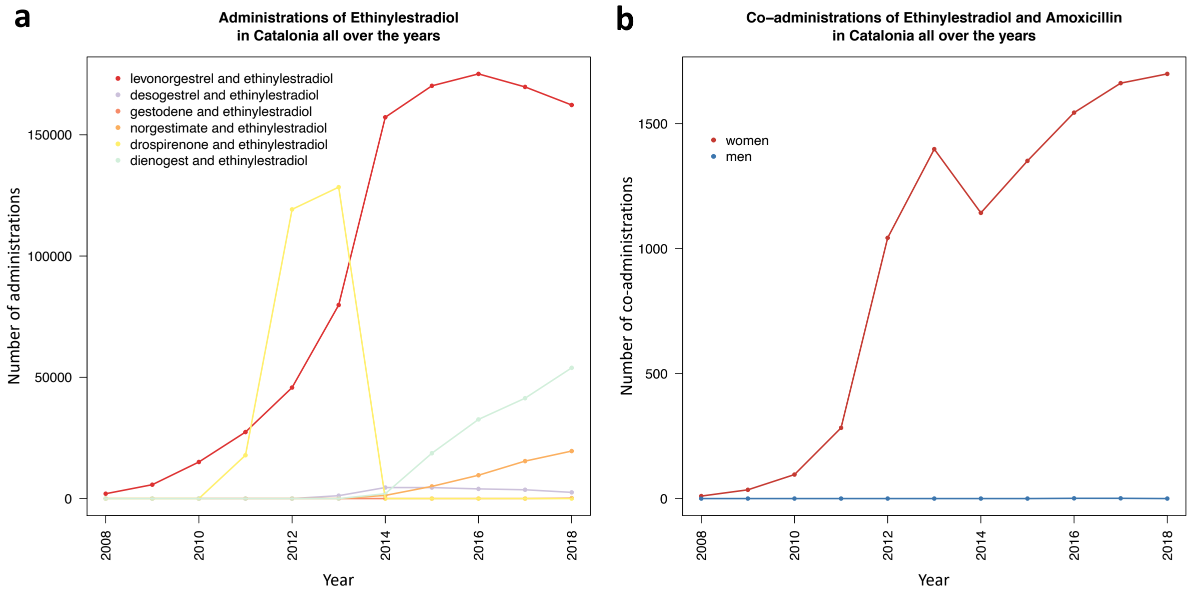


**Figure S7.** Evolution of the number of (a) administrations of Ethinylestradiol and (b) co-administrations of Ethinylestradiol and Amoxicillin from 2008 to 2018 in Catalonia. In b, red and blue colors denote the number of co-administrations in women and men respectively.

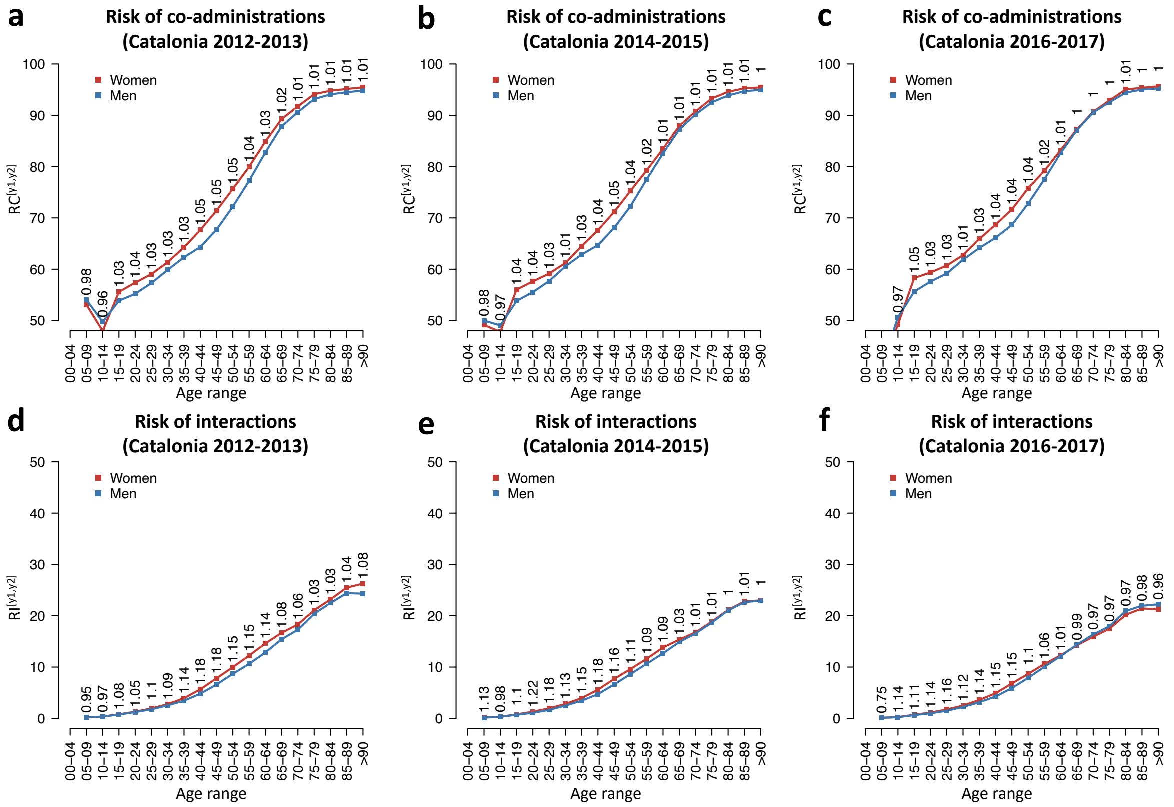

**Figure S8.** Risk of co-administration ($RC^{[y1,y2],g}$) and interaction ($RI^{[y1,y2],g}$) by age group ($[y1, y2]$) and sex ($g$) in Catalonia every two years after the removal of Ethinylestradiol dispensations. Red and blue colors denote the risk of co-administration in women and men, respectively. Relative risk of co-administration ($RRC^{[y1,y2],W}$) and interaction ($RRI^{[y1,y2],W}$) for women per age group shown above the points (as defined in sections 4.5 and 4.6).

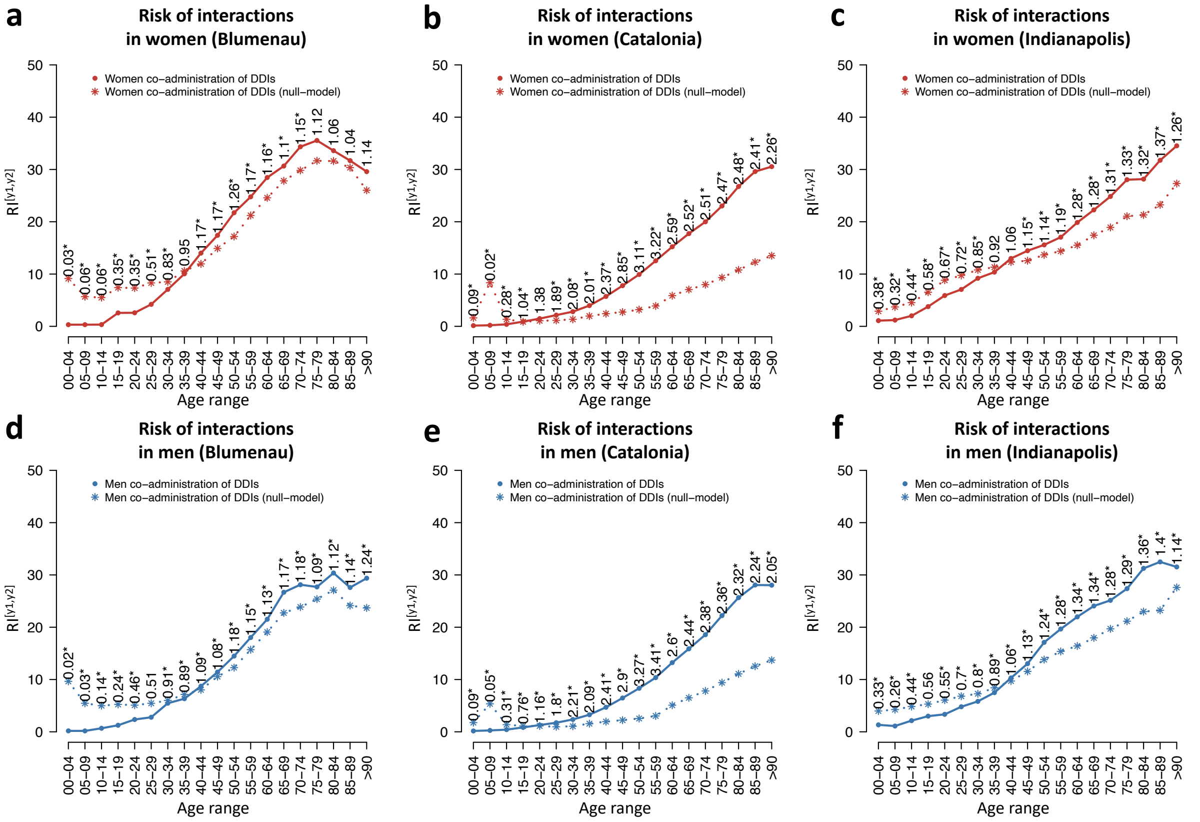


**Figure S9.** Risk of interaction ($RI^{[y1,y2],g}$) and its associated null model ($\widehat{RI}^{[y1,y2],g}$) by age group ($[y1,y2]$) and sex (*g*) in Blumenau, Catalonia, and Indianapolis during the first 18 months of administrations. Circles denote the values obtained with the real data ($RI$), while asterisks denote the values obtained using the null model ($\widehat{RI}$). Risk for women (*g* = *W*) shown in red (a-c) and men (*g* = *M*) shown in blue (d-f), respectively. The associated relative risk, $RI^{[y1,y2],g}/\widehat{RI}^{[y1,y2],g}$, shown over the points for each age group $[y1,y2]$.

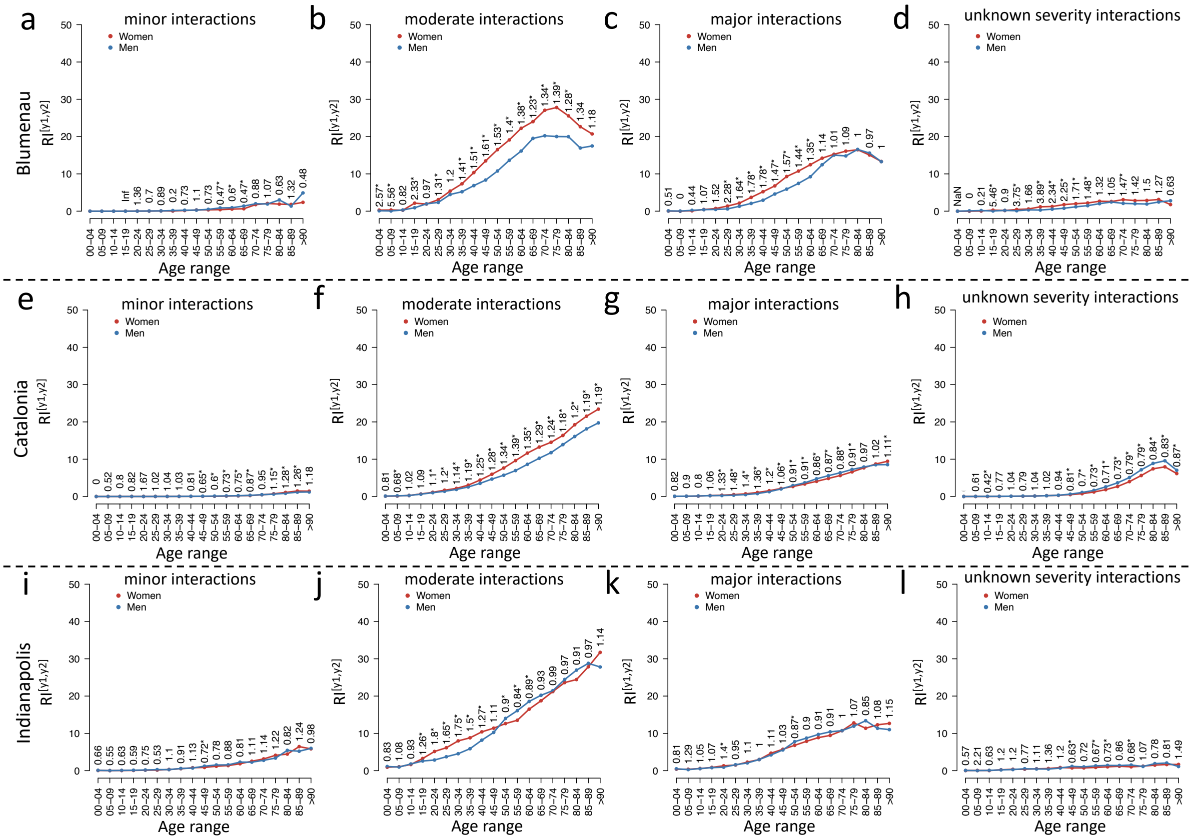


**Figure S10.** Risk of interaction ($RI^{[y1,y2],g}$) by severity in Blumenau (a-d), Catalonia (e-h), and Indianapolis (i-l) during the first 18 months of administrations. Red and blue colors denote the risk of co-administration in women and men respectively. The classification as minor (a, e, i), moderate (b, f, j), and major (c, g, k) interactions are extracted from drugs.com [16]. Those interactions with no description of the severity are denoted as "unknown severity interactions" (d, h, l). Relative risks of interaction ($RRI^{[y1,y2],W}$) for women per age group shown above points (as defined in sections 4.5 and 4.6). Asterisks denote significant differences (Fisher's exact test).

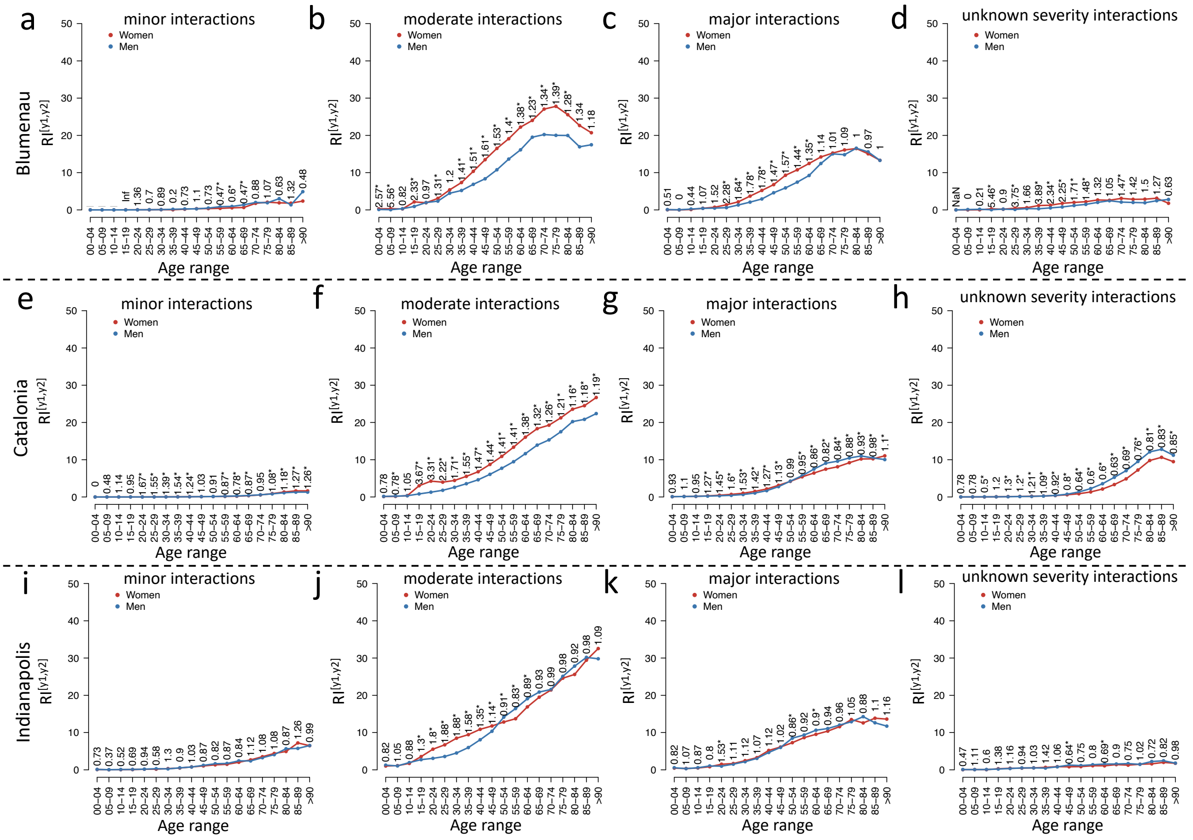


**Figure S11.** Risk of interactions ($RI^{[y1,y2],g}$) by severity score for Blumenau (a-d), Catalonia (e-h), and Indianapolis (i-l) during the whole study period for each population. Red and blue colors denote the risk of interaction for women and men, respectively. Severity score for minor (a, e, i), moderate (b, f, j), and major (c, g, k) interactions as per drugs.com [16]. Interactions with no associated severity score shown as "unknown severity interactions" (d, h, l). Relative risk of interaction ($RRI^{[y1,y2],W}$) for women per age group shown above points (as defined in sections 4.5 and 4.6). Asterisks denote significant differences (Fisher's exact test).

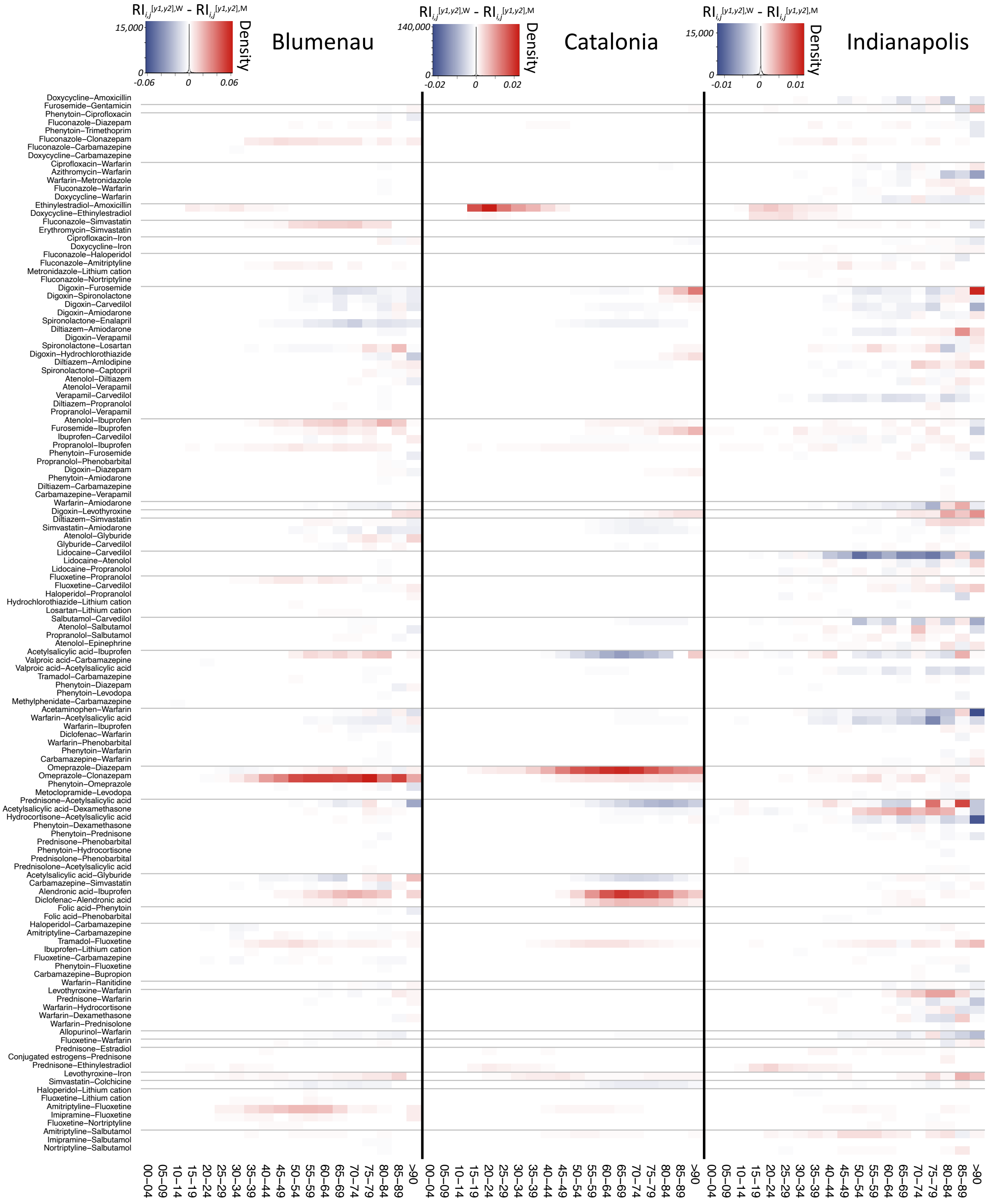

**Figure S12.** Sex-associated differential co-administration of the drug-drug interaction co-administration in the three populations with ageing. Heatmaps' colors denote the increased percentage of women (red) and men (blue) taking each DDI at each age bin in the different populations, calculated as $RI_{i,j}^{[y1,y2],W} - RI_{i,j}^{[y1,y2],M}$ (*eq. 11*).

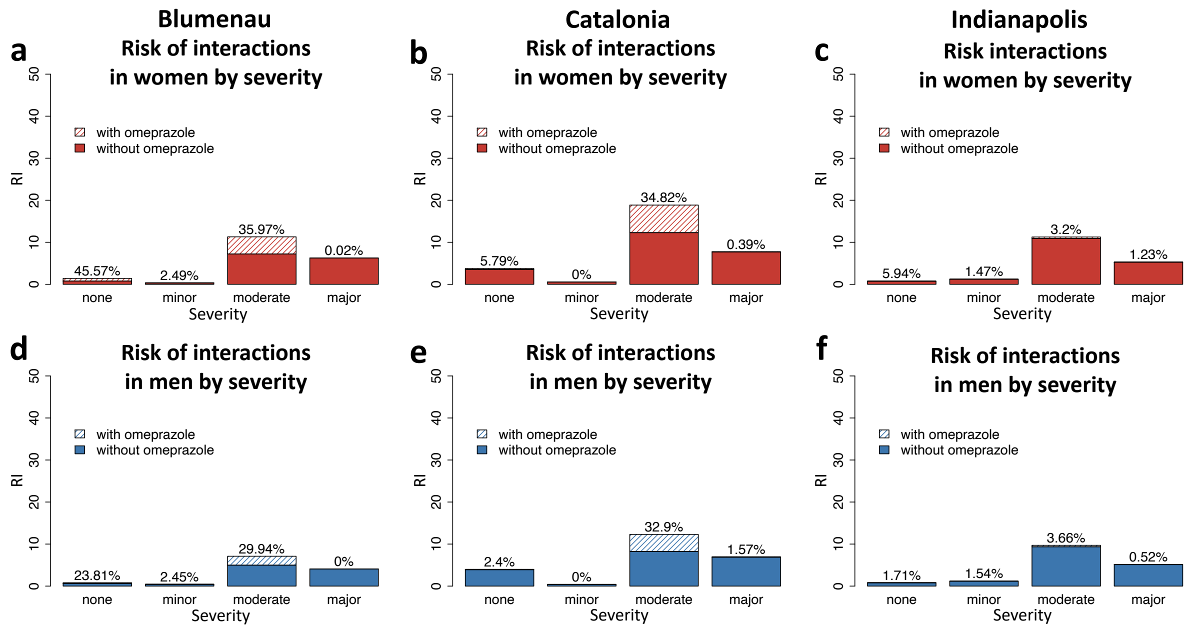


**Figure S13.** Risk of interaction (*RI*) by severity score before and after replacing Omeprazole with other PPIs (as defined in section 4.9).

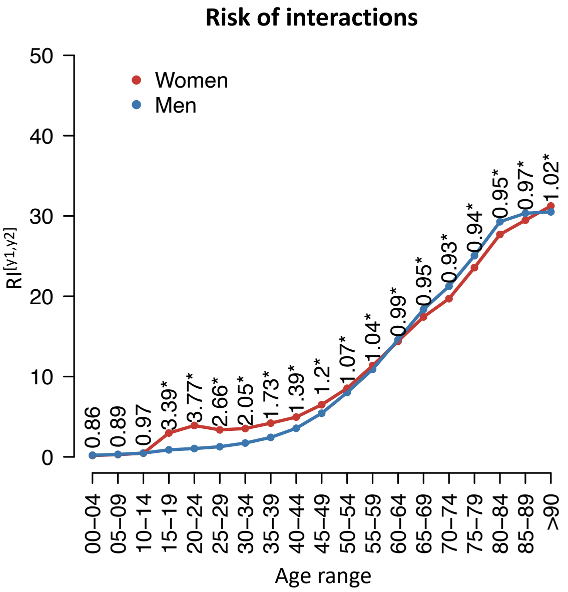


**Figure S14.** Risk of interaction by age and sex in Catalonia after the removal of Omeprazoleassociated interactions. Red and blue colors denote the risk of co-administration in women and men respectively. Relative risks of interaction $(RRI^{[y1,y2],W})$ for women per age group displayed above the points (as defined in sections 4.5 and 4.6). Asterisks denote significant differences (Fisher's exact test).

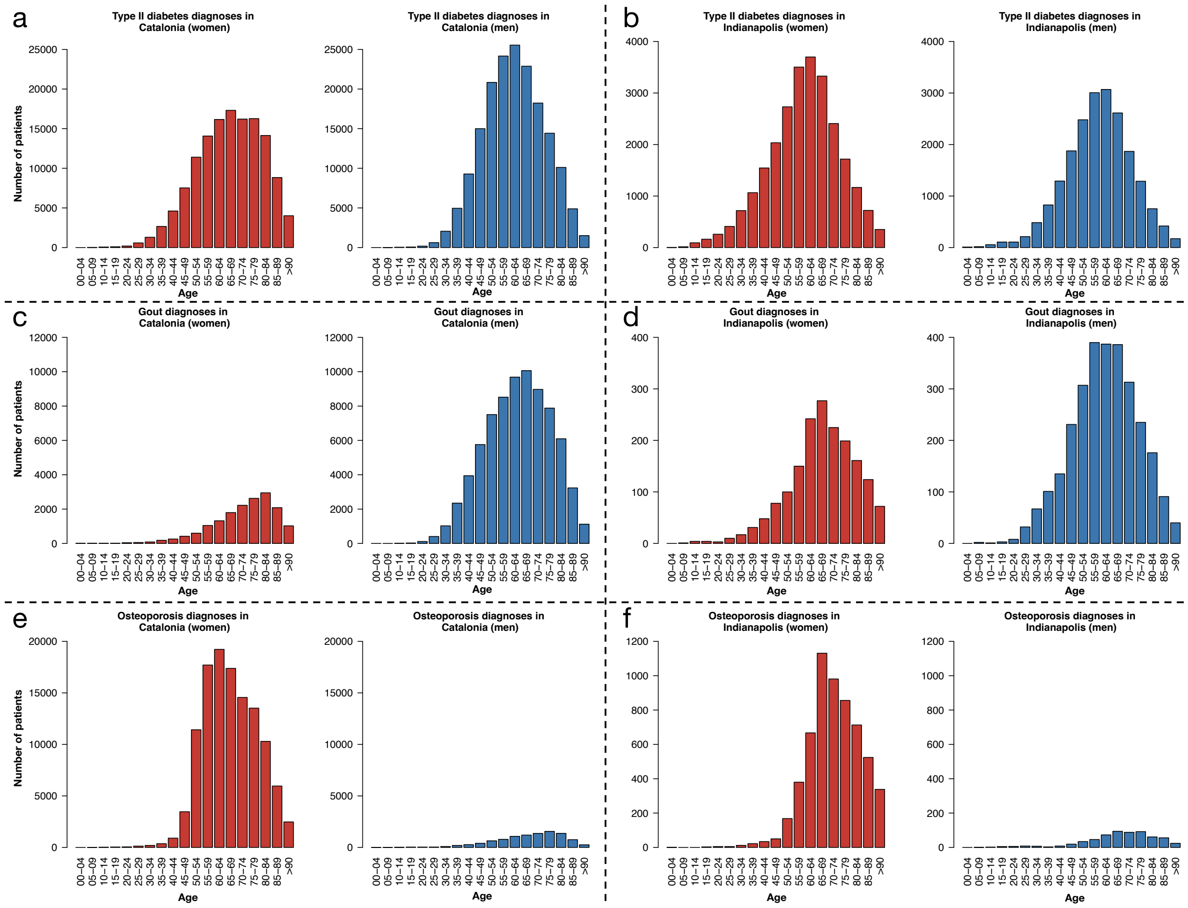


**Figure S15.** Type II diabetes, gout, and osteoporosis diagnoses in Catalonia and Indianapolis for women (red) and men (blue).

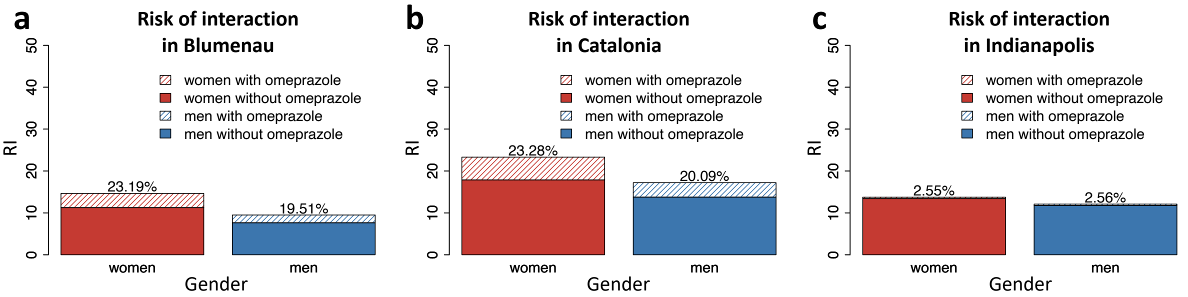


**Figure S16.** Risk of interaction (*RI*) before and after replacing Omeprazole with other protonpump inhibitors (PPI), as defined in sections 4.4 and 4.9.

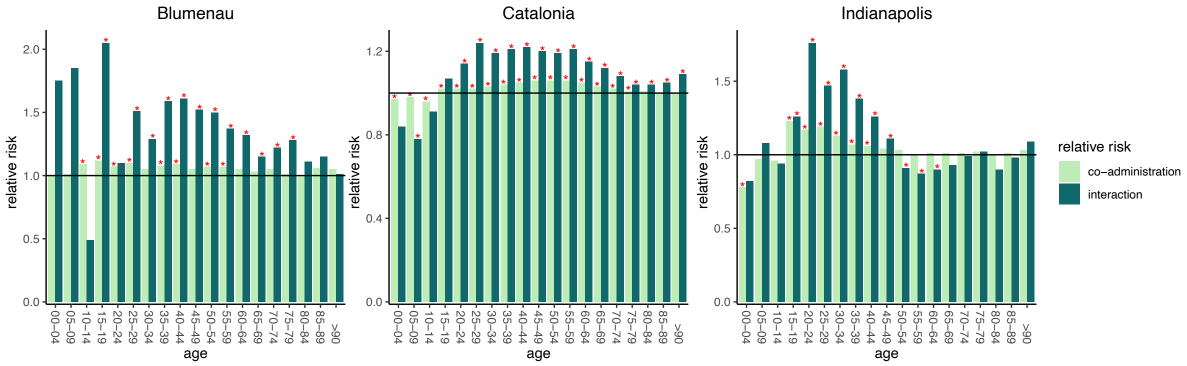


**Figure S17.** Relative risks of co-administration ($RRC^{[y1,y2],W}$) and interaction ($RRI^{[y1,y2],W}$) for women per age group (as defined in sections 4.5 and 4.6) for Blumenau, Catalonia, and Indianapolis in the first 18 months of administrations. Asterisks denote significant differences (Fisher's exact test). Note scales are different among plots.
